# Effectiveness of Homeopathic Interventions for Insomnia and Sleep Disorders: A Systematic Review and Meta-Analysis

**DOI:** 10.1101/2025.11.08.25339705

**Authors:** Muhammad Awais Paracha, Faryal Fazal, Sania Abdul Jalil Khan, Syeda Sidra Rizvi, Rabia Ali Afridi, Yasith Mathangasinghe

## Abstract

Insomnia is a common sleep disorder, and many individuals seek alternative treatments like homeopathy. However, evidence for its effectiveness remains controversial. This systematic review and meta-analysis evaluated the effectiveness of homeopathic interventions for insomnia and sleep-wake disorders. A comprehensive search of PubMed, MEDLINE, CINAHL, and the Cochrane Library was conducted for studies published between 2010 and 2025. We included randomized controlled trials (RCTs) and non-randomized studies involving adults (≥18 years) with primary insomnia receiving any homeopathic intervention compared to placebo, no treatment, or active care. Primary outcomes included validated sleep quality measures (e.g., Pittsburgh Sleep Quality Index (PSQI), Insomnia Severity Index (ISI). Four reviewers independently performed study selection, data extraction, and risk of bias assessment using RoB 2.0 and ROBINS-I. A random-effects meta-analysis was conducted for controlled trials, and a narrative synthesis for non-randomized studies. Certainty of evidence was assessed using Grading of Recommendations, Assessment, Development and Evaluation (GRADE). The search yielded 1304 records; 12 studies (nine RCTs and three non-randomized) met inclusion criteria. Meta-analysis showed a large, statistically significant positive effect of homeopathy on sleep outcomes (SMD = 0.81, 95% CI [0.24, 1.38], p = 0.0055), with substantial heterogeneity (I² = 86.04%) and publication bias (Egger’s test, p = 0.0079). Most studies had high or critical risk of bias, and overall certainty was low. Homeopathic interventions showed a large positive effect on sleep outcomes, but due to high bias, heterogeneity, and publication bias, evidence remains low-certainty and insufficient to support effectiveness. High-quality RCTs are needed.

**Systematic Review Registration:** PROSPERO CRD42025649926.

## Introduction

According to the American Psychiatric Association (APA), insomnia is the most prevalent sleep disorder worldwide and is characterized by difficulty in initiating or maintaining sleep, early-morning awakening and an inability to return to sleep, occurring for at least three nights a week over a three-month period [1]. Insomnia impairs daytime functioning and can be caused by a variety of factors [1]. It frequently co-occurs with underlying medical conditions such as cardiovascular diseases, renal disorders, or obstructive airway diseases [2]. The long-term use of certain medications, psychological disorders, substance use disorder (SUD) aging, genetic predisposition, or traumatic brain injuries can also contribute to its development [3]. A systematic review report that 52.57% of the global population experienced sub-threshold insomnia, while 13.75% suffer from moderately severe clinical sleeplessness, and approximately 2.5% have severe clinical insomnia [4]. In the United States, it affects approximately 10% of the population [5]. If left untreated, chronic insomnia can significantly impact an individual’s economic, social, and occupational well-being [1].

Disturbed sleep not only impairs daytime performance, alertness, and concentration, but is also associated with an increased risk of depression and other psychiatric disorders. For instance, individuals who experienced insomnia for two weeks or longer had a higher risk of developing depression over time [6]. Many individuals resort to pharmacological treatments; however, these medications can lead to dependency, making it difficult for patients to sleep without them [7]. Self-medication is common but can be dangerous, as some frequently used options, such as antihistamines, are not recommended for insomnia due to their adverse effects [8]. These adverse effects can include fatigue and light-headedness, and, in the long run, may lead to cognitive impairment and memory loss. Furthermore, discontinuing these medications often results in the recurrence of insomnia.

Due to the adverse effects of pharmacological interventions, many people seek alternative treatments, such as homeopathy, for managing insomnia [9]. Homeopathy is a patient-centred treatment where dosage can vary, contributing to uncertainty in outcomes [10]. Based on the principle that “like cures like”, homeopathy utilizes extremely diluted substances intended to stimulate the natural healing processes of the body. Potential mechanisms through which it may influence sleep include the modulation of stress responses and the regulation of circadian rhythms. However, the individualized nature of homeopathic prescriptions, variations in remedy formulations, and differences in patient characteristics add complexity to its research and interpretation of findings. The evidence supporting homeopathic treatments for insomnia is mixed; while some placebo-controlled trials show encouraging results, others have failed to demonstrate significant advantages over placebo [11, 12]. This variability combined with methodological issues such as a lack of randomized controlled trials, small sample sizes, the absence of standardized outcome measures, and variations in treatment protocols—calls the efficacy of homeopathy for insomnia into question [13].

An earlier systematic on this topic was conducted by Cooper and Relton in 2010 found no statistically significant effect of homeopathic interventions on insomnia [14]. Since its publication, several developments have justified an updated review. A subsequent systematic review by Ernst in 2011 also found no significant differences between homeopathy and placebo for insomnia or sleep disorders [15]. More recent homeopathy-focused reviews have also emerged, but they fail to address the on-going uncertainty due to their methodological limitations [16, 17]. For instance, Sreevidhy et al, (2023) review [16] is a descriptive literature review that lists a few homeopathic remedies for insomnia and does not fulfils the definition of a systematic review [18]. Similarly Aphale, Sharma and Shekhar 2024 [17] review also do not follow rigorous methods in their systematic review such as risk of bias assessment but they found potential benefits of homeopathy for insomnia and raises a controversy [19]. These reviews identify a gap that justices a rigorous systematic review of the existing literature. Additionally, the emergence of new trials, alongside advances in trial design and appraisal methods, necessitates a re-evaluation of recent evidence. An updated systematic review, conducted within a rigorous contemporary framework using tools such as the Preferred Reporting Items for Systematic Reviews and Meta-Analyses (PRISMA) 2020, the Risk of Bias 2.0 (RoB 2.0) tool, and the Grading of Recommendations, Assessment, Development, and Evaluations (GRADE) approach [19, 20, 21] is required to more precisely estimate effects and assess the effectiveness of homeopathy for managing insomnia.

This review aims to investigate the types of homeopathic interventions used for insomnia and to evaluate the existing evidence on their effectiveness for insomnia and sleep-wake disorders. The review will explore whether homeopathic therapies, when compared to placebo or conventional treatments, can improve sleep quality, duration, and other related health outcomes in individuals with insomnia and sleep disorders.

## Methods

### Registration and Protocol

The protocol for this systematic review was registered with PROSPERO (International Prospective Register of Systematic Reviews) under the registration number CRD42025649926, available from: https://www.crd.york.ac.uk/prospero/display_record.php?ID=CRD42025649926.

We strictly followed the registered protocol, and there were no deviations to report. However, several modifications were made to the initial protocol to minimize the risk of bias and enhance clarity. All revisions were documented in an updated version on PROSPERO before the data extraction stage. The title was modified from “Sleep-related Disorders” to “Sleep Disorders,” and a corresponding change was made to the objectives and eligibility criteria. We also added the keyword “Sleep-Wake Disorders.” To minimize language bias, the explicit exclusion of non-English articles was removed from the search strategy. The inclusion criterion “observational studies without a control group” was broadened to “observational studies.” We added exclusion criteria for studies where the full text was unavailable after institutional access and author contact attempts, and clarified that non-English studies would be excluded only if a translation could not be obtained using Google Translate or AI tools. The eligibility criteria were further refined to include “only primary insomnia populations.” We also modified the order of the primary and secondary outcomes, clarified that manual screening would be conducted “without AI-assisted ranking,” updated the risk of bias tool from RoB 1.0 to RoB 2.0, specified the order of outcome prioritisation for data synthesis (“PSQI global score is prioritised; if unavailable, Epworth Sleepiness Scale (ESS) will be used, followed by sleep latency”), and added a plan to “conduct pre-specified subgroup analyses based on clinically relevant factors.”

### Eligibility Criteria

Eligibility criteria were defined using the Population, Intervention, Comparator, and Outcomes (PICO) framework:

- Population (P): Studies including adults (aged 18 years or older) with a diagnosis of primary insomnia were eligible.
- Intervention (I): Any homeopathic treatment, including single or combination medicines administered via any route.
- Comparator (C): Placebo, no treatment, or routine active care (e.g., cognitive behavioural therapy for insomnia [CBT-I] or pharmacological treatment). Studies evaluating only allopathic or herbal interventions (i.e., without a homeopathy arm) were excluded.
- Outcomes (O): Primary outcomes included sleep quality measured with validated tools such as the Pittsburgh Sleep Quality Index (PSQI), Jenkins Sleep Scale (JSS), Insomnia Severity Index (ISI), or Sleep Impairment Index (SII), as well as self-reported sleep duration and latency from baseline to final follow-up. When multiple outcomes were reported, the PSQI global score was prioritized as the primary outcome. Secondary outcomes included the frequency of nighttime awakenings, sleep satisfaction, total sleep time, adverse events, quality of life, and changes in related health conditions like anxiety or depression.

### Information Sources

We conducted a comprehensive search for studies published between 2010 and 2025. The search covered a broad range of bibliographic databases, including PubMed, MEDLINE, CINAHL EBSCO, the Cochrane Library, Euro PMC, and the Directory of Open Access Journals. We also searched clinical trial registries, such as ClinicalTrials.gov and the World Health Organization International Clinical Trials Registry Platform. To ensure thorough coverage and minimize publication bias, the search was expanded to include grey literature sources like Google Scholar, dissertations and theses from Open Access Theses and Dissertations and ProQuest, and the reference lists of included studies and relevant systematic reviews.

The search strategy combined keywords and medical subject headings (MeSH terms), which were adapted for each database using Boolean operators. For instance, the PubMed search string was: ((“insomnia”[MeSH] OR “sleep initiation and maintenance disorders”[MeSH] OR “insomnia”[Title/Abstract]) AND (“homeopathy”[MeSH] OR “homeopathic”[Title/Abstract])). The complete search queries for all databases are detailed in the Appendix I: Search Strategy.

### Selection Process

The study selection process was conducted in two distinct phases. In the first phase, titles and abstracts were screened using the Rayyan web application [22]. In the second phase, the full texts of potentially relevant articles were retrieved and carefully assessed against the eligibility criteria. The reasons for excluding studies were documented at both the screening and full-text review stages. To ensure reliability, four reviewers (MAP, FF, SAJ, and SSR) independently screened all titles and abstracts in a double-blinded fashion. Any disagreements were resolved through discussion among the reviewers or, when necessary, with the involvement of a fifth reviewer (RAA). This same rigorous process was applied during the full-text review. For quality control, the review team also re-screened a random selection of the articles.

### Data Collection Process

Data were independently extracted by four reviewers using a standardized Excel spread sheet. This data extraction form was adapted from the Cochrane data collection form [18] and incorporated elements from the Template for Intervention Description and Replication (TIDieR) checklist [23] to ensure a thorough description of the interventions. The form was piloted before use. For one study Harrison, (2011) [33], standard deviations were not reported, and the author could not be contacted; in this instance, missing values were imputed using an available-case analysis. To maintain accuracy and completeness, every entry in the data extraction sheet was cross-verified by an experienced team member (MAP). We used a structured form to extract data on: (1) Study Characteristics, including the study identifier, authors, publication year, country, setting, design, and sample size; (2) Participant Demographics, such as age, sex, and baseline characteristics; (3) Intervention and Comparator Details, including the name of the homeopathic medicine, its potency, dosage, frequency, treatment duration, and the provider; and (4) Outcomes, including all reported primary and secondary outcomes measured at baseline and all follow-up points.

### Risk of Bias Assessment

We assessed the risk of bias for non-randomized and randomized studies separately using pre-specified, validated instruments. Four reviewers (MAP, SAJ, SR, FF, and RAA) individually evaluated each domain within each study. The assessments were conducted in duplicate with a shuffled reviewer order; any disagreements were resolved by consensus.

### Randomized Controlled Trials

We assessed the risk of bias for included studies using the revised Cochrane Risk of Bias tool for randomized trials (RoB 2.0), with additional guidance for crossover trials where appropriate [19]. RoB 2.0 covers five domains: bias arising from the randomization process; bias due to deviations from intended interventions; bias due to missing outcome data; bias in the measurement of the outcome; and bias in the selection of the reported result. Before beginning the assessments, the entire RoB 2.0 guidance document was read thoroughly and followed to ensure a consistent and transparent process. For each domain, we answered signalling questions provided by the tool to gather relevant information. These questions were answered with the recommended standard response options (“Yes,” “Probably yes,” “Probably no,” “No,” and “No information”). Our judgment for each domain was then determined directly from these responses using the decision algorithms integrated into the RoB 2.0 framework. For accuracy and transparency, we extracted verbatim text from the studies to use as supporting statements for each signalling question. This practice allowed us to clearly record the rationale behind each response, reducing the potential for subjective decisions. For example, if a study explicitly described its randomization or blinding technique, we quoted the original description as a supporting statement. When studies did not explicitly report but implied methodological decisions (e.g., an intention-to-treat analysis), this was recorded as “Y (ITT implied).”

We recorded all judgments and supporting statements in the official MS Excel-based RoB 2.0 tool downloaded from the Cochrane risk of bias website (https://www.riskofbias.info/welcome/rob-2-0-tool/current-version-of-rob-2). This macro-enabled form facilitates structured data entry, generates summary tables, and produces traffic-light figures to visually present the distribution of risk-of-bias judgments across studies and domains. Two reviewers independently applied the RoB 2.0 tool to each included study, documenting supporting information and justifications for each domain’s risk of bias judgment (low risk, some concerns, or high risk). Discrepancies in judgments were resolved through discussion, and a third reviewer was available to arbitrate if a consensus could not be reached.

### Non-Randomized Controlled Trials

We also conducted a robust risk of bias assessment for non-randomized studies using the ROBINS-I tool, which assesses seven key domains: bias due to confounding, bias in the selection of participants, bias in the classification of interventions, bias due to deviations from intended interventions, bias due to missing data, bias in the measurement of outcomes, and bias in the selection of the reported result [24]. Two reviewers independently applied the instrument, extracting verbatim text from the publications to justify their judgments. Each domain was assessed using signalling questions to reach a judgment on the risk of bias (low, moderate, serious, or critical) consistent with the ROBINS-I algorithms. Any disputes were settled through consensus discussion or third-reviewer arbitration. The results were summarized in an MS Excel sheet that incorporated direct quotations from the studies as supporting text. We produced traffic-light plots and weighted bar charts to summarize the risk of bias across studies using the Risk of Bias Visualization (robvis) software [25]. This process allowed for a transparent assessment of potential biases, such as residual confounding, attrition bias, and detection bias in unblinded comparisons, with each judgment explicitly linked to textual evidence.

### Effect Measures

All data processing and statistical analyses were performed in R using the metafor, tidyverse, and readxl packages [26]. For each eligible study, we extracted the mean, standard deviation (SD), and sample size (N) for both the intervention and control arms.

### Effect Size Calculation

The primary outcome was the change in sleep-related measures. The effect size for each study was calculated as the standardized mean difference (SMD) using Hedges’ g to correct for small sample bias. To ensure a consistent direction of effect, where a positive SMD always indicates a favorable outcome for the intervention, the signs of SMDs from studies using ‘reverse’ scales (e.g., the Pittsburgh Sleep Quality Index [PSQI], where lower scores are better) were mathematically inverted prior to analysis. Before calculating the effect size, outcomes measured in hours or weeks were standardized to minutes or days, respectively.

### Synthesis Methods

The analysis employed a mixed-methods approach for evidence synthesis. To evaluate effectiveness, we conducted a meta-analysis of the randomized trials using random-effects models and assessed statistical heterogeneity using the I² statistic. For the non-randomized or observational studies, we conducted a narrative synthesis.

### Narrative Synthesis

A narrative synthesis was conducted to analyse and interpret the results of the non-randomized trials included in the review. The synthesis followed a systematic, multi-phased approach to minimize bias and maximize transparency. First, we tabulated the study characteristics and findings to identify emerging patterns. Next, the studies were categorized based on common characteristics, such as study design, to facilitate comparison. We then analysed relationships within and across studies to explore potential determinants of variability in the direction and magnitude of effects, considering elements such as intervention complexity, mode of delivery, and risk of bias. This systematic process enabled a structured analysis of the data, allowing the review to evaluate the efficacy of homeopathy interventions for sleep disorders.

### Meta-Analysis

To synthesize the findings and calculate an overall effect from the randomized trials, we performed a random-effects meta-analysis using the Restricted Maximum-Likelihood (REML) estimator. This approach provides a single summary effect while accounting for both within-study sampling error and between-study heterogeneity.

### Heterogeneity and Publication Bias Assessment

The degree of between-study heterogeneity was quantified using the I² statistic, which describes the percentage of total variation across studies attributable to true differences rather than chance. The statistical significance of this heterogeneity was assessed with Cochran’s Q test. Potential publication bias was evaluated visually by inspecting a funnel plot and statistically using Egger’s regression test for funnel plot asymmetry. A p-value < 0.05 was considered statistically significant.

### Reporting Bias Assessment

Publication bias for the randomized controlled trials was assessed using funnel plots and Egger’s regression test.

### Certainty Assessment

The certainty of the evidence was evaluated independently by two reviewers (MAP, SAJ). We graded the evidence using GRADEpro GDT (gradepro.org), following Cochrane’s GRADE guidance [27, 28]. Following the GRADE framework, we examined five key factors to determine the certainty of the evidence for the predefined outcomes: study limitations, consistency of effect, imprecision, indirectness, and publication bias [29]. The certainty was classified as high, moderate, low, or very low. Where applicable, we also considered reasons to upgrade or downgrade the certainty rating based on criteria such as a large effect size, a dose-response relationship, or plausible confounding influences. All decisions to adjust the certainty rating, whether downgrading or upgrading, were documented with explanatory footnotes.

## Results

### Study Selection

The initial search of databases and registries, conducted from March to May 2025, identified 1,304 records. After removing 317 duplicates, 987 unique records advanced to the title and abstract screening stage. At this stage, 920 articles were excluded. Sixty-seven studies proceeded to a full-text screening, where 56 were excluded for the following reasons: background article (n=2), ineligible population (n=4), ineligible outcome (n=3), ineligible intervention (n=15), ineligible study design (n=7), ineligible publication type (n=19), and ineligible study duration (n=16). Ultimately, 11 studies met the eligibility criteria and were included in the final analysis. An additional study, identified later through a Google search, also met the eligibility criteria, bringing the total number of included studies to 12. The study selection process, which followed the Preferred Reporting Items for Systematic Reviews and Meta-Analyses (PRISMA) methodology [18], is illustrated in Figure 1: Appendix II

### Study Characteristics

The characteristics of the 12 included studies are summarized in Table 1: Appendix III: Study Characteristics. The studies included randomized controlled trials (RCTs) and observational studies with pre-post-test designs. These studies were conducted in India, South Africa, France, and Germany between 2010 and 2025. Participant ages ranged from 18 to 91 years, with sample sizes varying from 28 to 639.

Interventions consisted of individualized homeopathic medicines (6 studies), specific preparations such as Eschscholzia californica (1 study) and Passiflora incarnata (1 study), and combination remedies including Dysto-loges S (1 study), Passiflora Composé (1 study), a homeopathic complex (1 study), and Coffea cruda 200cH (1 study). Comparators included placebo, placebo combined with remedial measures, and Passiflora Composé combined with complementary therapies. Intervention durations ranged from two weeks to four months. Most studies measured outcomes at multiple time points, such as at baseline, during follow-up, and at the end of treatment (e.g., at two, four, or six weeks). The shortest follow-up period was 10 days, while the longest extended to four months. Some studies conducted frequent assessments (e.g., every 15 days or weekly), whereas others measured outcomes only at baseline and the final intervention point. The most frequently prescribed individualized homeopathic remedies were Nux vomica, Pulsatilla, Natrum muriaticum, Sulphur, Coffea cruda, Lachesis, Phosphorus, and Ignatia, each appearing in 25% to 40% of the individualized intervention studies. Standardized preparations primarily featured Passiflora incarnata (used in 25% of all studies), followed by Eschscholzia californica and other complex formulations. A detailed description of each intervention is provided in Table 2: Appendix II: Intervention Characteristics.

Sleep-related outcomes were primarily measured using validated tools. The Pittsburgh Sleep Quality Index (PSQI) was used in five studies (41.7%), the Insomnia Severity Index (ISI) in three (25%), the Jenkins Sleep Scale (JSS) in one (8.3%), and the Visual Analogue Scale for Sleep (VISS) in one (8.3%). General self-reported sleep quality ratings were used in two studies (16.7%), and sleep diaries were used in four (33.3%). Placebo controls were used in 33% of the studies, while 17% compared different homeopathic regimens or combined treatments. The remaining 25% were observational studies without a control group. Dropout rates were generally low; approximately 58% of studies reported between zero and six dropouts. However, one study (8%) had significantly higher attrition, with 25 participants lost to follow-up, and 33% of studies did not report dropout details.

All interventions were administered orally as drops, tablets, globules, or sachets. The majority of interventions (over 60%) were delivered by qualified homeopathic physicians or practitioners, with the remainder provided by general practitioners (20%) or research organizations and independent dispensers (20%). The primary study setting was outpatient departments (approximately 75%), with other studies conducted in university clinics, private practices, and general practitioner offices. Nearly two-thirds of the studies reported protocol modifications, particularly in individualized prescriptions where the remedy and potency were adjusted; the remaining studies used fixed-dose regimens. Adherence and fidelity were reported in less than half of the studies (approximately 40%), typically monitored through follow-up appointments and sleep diaries (Table 3: Appendix III: TIDier Checklist for Interventions).

### Risk of Bias

We assessed the nine randomized trials using the Cochrane Risk of Bias tool 2.0 (RoB 2.0), which includes specific guidance for parallel-arm and crossover designs. The overall risk of bias was judged to be high in seven trials, low in one and of some concern in one. Figure 2 Appendix II: Risk of Bias Assessment (Parallel Arm RCTs) shows an overview of the judgment distribution per domain.

Seven trials were rated at a high overall risk of bias: Kolia-Adam 2010 [30], Arora 2019 [31], Sakshi 2024 [32], Harrison 2011 [33], Prajakta et al. 2024 [34], and Hellhammer and Schubert 2013 [35]. The trial by Naudé et al. 2010 [36] was judged to have some concerns, and only the study by Michael et al. 2019 [37] was rated at a low risk of bias across all domains.

Issues in Domain 1 (randomization process) were common, often due to unclear randomization procedures or insufficient reporting of allocation concealment. Kolia-Adam 2010 [30] and Sakshi 2024 [32] were rated at high risk in this domain, while Arora 2019 [31], Harrison 2011 [33], and Prajakta 2024 [34] had some concerns. In Domain 2 (deviations from intended interventions), nearly half of the studies were rated at low risk. However, Sakshi 2024 [33] and Harrison 2011 [33] were rated at high risk due to a lack of clarity regarding blinding and deviations from the intended interventions. In Domain 3 (missing outcome data), Sakshi 2024 [32], Harrison 2011 [33], and Prajakta 2024 [34] were rated at high risk because dropout patterns may have biased the results. Naudé et al. 2010 [36] raised some concerns due to incomplete reporting of attrition. Five studies were rated at low risk in Domain 4 (measurement of the outcome). In contrast, Kolia-Adam 2010 [30], Arora 2019 [31], and Sakshi 2024 [32] were rated at high risk because outcome assessors were not blinded or the measures were potentially influenced by knowledge of the intervention. Domain 5 (selection of the reported result) revealed a consistent issue with selective reporting or incomplete outcome data. The studies by Kolia-Adam 2010 [30], Arora 2019 [31], Sakshi 2024 [32], Harrison 2011 [33], and Hellhammer and Schubert 2013 [35] were rated at high risk, while Prajakta 2024 [34] and Naudé et al. 2010 [36] raised some concerns. In summary, only the study by Michael et al. 2019 [37] was judged to have a low risk of bias across all domains, suggesting it provides the most robust evidence among the included RCTs.

The crossover trial by Motiwala (2022) [38] was assessed to be at an overall high risk of bias. Concerns were raised regarding the randomization process (Domain 1) due to inadequate information on allocation concealment. A high risk was identified for bias arising from period and carryover effects, as well as for deviations from the intended intervention, reflecting potential residual effects and a lack of blinding. While missing outcome data (Domain 3) was rated as low risk, measurement of the outcome (Domain 4) raised some concerns, and selection of the reported result (Domain 5) was rated as high risk due to the absence of a pre-specified analysis plan.

We also evaluated the risk of bias for three non-randomized studies (Villet et al. 2015 [39], Parmar et al. 2025 [40], and Singh et al. 2021 [41]) using the ROBINS-I (Risk Of Bias In Non-randomized Studies of Interventions) assessment tool. There was a critical risk of bias in Domain 1 (confounding) related to uncontrolled factors, as all studies used a single-arm pre-post design with no control group. Domain 2 (intervention classification) was classified as a moderate risk because of the risk of misclassification, and Domain 3 (participant selection) was a low risk because the studies incorporated a new-user design. Domain 4 (deviations from intervention) was of low-to-moderate risk, as the studies were observational. Domain 5 (missing data) varied from a serious risk (Villet et al. 2015 [39], Parmar et al. 2025 [40]) due to complete-case analysis, to a low risk (Singh et al. 2021 [41]), where no data were missing. Domain 6 (outcome measurement) posed a serious risk because outcome assessors were not blinded, and Domain 7 (selective reporting) was a moderate risk due to the failure to report pre-specified analysis plans. Overall, all three non-randomized studies were judged to be at a critical risk of bias. The absence of control groups and blinding likely introduced a bias in favour of the intervention, making it impossible to separate true treatment effects from confounding.

## RESULTS OF INDIVIDUAL STUDIES

The included studies assessed a range of sleep-related outcomes across different time points using validated tools. Most studies that compared an intervention to a control group found favourable outcomes for the intervention. For example, some studies [34, 37, 38] reported significantly lower PSQI or ISI scores in the intervention arm, indicating improved sleep quality or reduced insomnia severity. Others reported increased sleep duration [30, 31, 36] or improved subjective sleep quality [35]. The single-arm observational studies also reported improvements in sleep outcomes post-intervention [39, 40, 41]. Overall, the results consistently suggest a positive impact of the interventions on sleep quality and duration across diverse measures and follow-up periods.

## RESULTS OF SYNTHESES

The narrative synthesis of findings of three observational studies i.e. Villet et al. (2015) [39], Parmar et al. (2024) [40], and Singh et al. (2021) [41], identified a consistent pattern of statistically significant improvement in sleep outcomes following homeopathic interventions despite variations in methodology and measurement. For instance, Villet et al. (2015) [39] conducted an open-label, observational study to investigate the homeopathic medicine Passiflora Composé (PC) in a primary care setting. The study enrolled 639 individuals, the majority of whom were female (78.6%), with a high prevalence of comorbid sleep disorders (74.0%), as assessed by the Jenkins Sleep Scale (JSS). The average baseline JSS score was 15.24 (±5.28). After four weeks of treatment, there was a statistically significant improvement in sleep scores (p < 0.001). The mean JSS score dropped by 4.48 points (a 29.4% reduction from baseline) to a final score of 10.76 (±4.67). Clinically, 53.9% of the patient group (n=366) experienced improved sleep, 40.1% remained stable, and 6.0% deteriorated.

Similarly, Parmar et al. (2024) [40] conducted a prospective, single-arm interventional study to evaluate the effectiveness of personalized homeopathic interventions for insomnia. The trial included 40 patients diagnosed with insomnia according to DSM-5 criteria, with outcomes measured using the Insomnia Severity Index (ISI). After three months of treatment, a paired t-test of the intention-to-treat sample (n=40) revealed a clinically significant improvement. Specifically, a majority of patients showed marked improvement (n=23; 57.5%), while another 35% (n=14) showed moderate improvement. Overall, 92.5% of the cohort experienced some degree of benefit; only three patients (7.5%) reported no improvement.

The third study, by Singh et al. (2021) [41], was also a prospective observational study, conducted in a specialized geriatric population (aged 60 and above) with insomnia. The study recruited 30 patients from the outpatient department of a homeopathic hospital. The primary outcome was the change in the global score of the Pittsburgh Sleep Quality Index (PSQI). The findings showed a statistically significant improvement after homeopathic treatment, with a paired-samples t-test revealing a mean reduction in the PSQI score of 3.66 points (t(29) = 7.67, p = 0.005).

Therefore, despite methodological heterogeneity including differences in study design, sample size, specific homeopathic interventions, patient populations, and the sleep scales used (JSS, ISI, and PSQI) a consistent result was found across all three studies. All studies reported a statistically significant improvement in sleep outcomes following homeopathic treatment. These improvements were observed in diverse populations, including a large female sample with comorbid anxiety and a specialized geriatric cohort, with a significant number of patients in all studies exhibiting improved sleep outcomes.

### Overall Meta-Analysis

A total of nine studies providing data for a controlled comparison were included in the final meta-analysis. Three of the nine outcomes were measured on ‘reverse’ scales, and their effect sizes were inverted accordingly to ensure a consistent direction of effect. The random-effects meta-analysis, combining all nine studies, yielded a pooled standardized mean difference of 0.81 (95% CI [0.24, 1.38]). This overall effect was statistically significant (p = 0.0055), indicating a positive impact of the interventions on sleep outcomes. The analysis revealed substantial and statistically significant heterogeneity across the studies (Q(df = 8) = 56.72, p < 0.0001; I² = 86.04%). The forest plot (Figure 8) displays the effect size for each individual study along with the overall pooled summary estimate

### Publication Bias

The Egger’s regression test for funnel plot asymmetry indicated a statistically significant presence of publication bias (t = 3.68, df = 7, p = 0.0079). The funnel plot (Figure 9) visually supports this finding, showing asymmetry, which suggests that smaller studies with non-significant or negative findings may be underrepresented.

The funnel plot displays the standardized mean difference (SMD) against the standard error for each study included in the meta-analysis. The dashed vertical line represents the overall pooled SMD, and the dashed diagonal lines indicate the 95% confidence limits in the absence of heterogeneity and publication bias. Asymmetry in the plot can suggest the presence of publication bias, where smaller studies with non-significant or negative findings may be underrepresented.

### Certainty of Evidence

The GRADE approach was used to assess the certainty of the evidence for each critical outcome [28]. The findings indicate that, compared to a placebo, homeopathy has a large effect on sleep outcomes (SMD = 0.81). However, the certainty of this evidence is low. The certainty rating was downgraded primarily due to serious concerns about the overall risk of bias across the included studies and the potential for publication bias. A low certainty rating means that the true effect may be substantially different from the estimated effect. A detailed breakdown of the GRADE assessment is provided in the footnotes of the GRADE Summary of Findings (Table 5: Appendix III).

## Discussion

Our systematic review and meta-analysis sought to consolidate evidence from randomized controlled trials on the efficacy of homeopathic interventions for improving sleep outcomes, specifically for insomnia. The meta-analysis of nine controlled trials found a significant positive effect of homeopathic interventions on sleep outcomes, with a pooled standardized mean difference (SMD) of 0.81 (95% CI 0.24–1.38) [30, 31, 32, 33, 34, 35, 36, 37, 38]. The observed pooled effect is broadly consistent with the individual findings of our narrative synthesis and several other trials. For example, a randomized controlled trial by Jong et al. (2016) involving 179 children with sleep disorders reported moderate-to-large improvements in insomnia symptoms compared with placebo; however, both the intervention and control groups improved significantly, which highlights the strong placebo response often seen in sleep disorder trials [42]. Similarly, Frass et al. (2020) conducted a prospective, randomized, placebo-controlled trial to investigate whether homeopathic treatment as an add-on therapy in non-small cell lung cancer patients could improve overall quality of life, and the authors also found benefits for secondary sleep outcomes [43]. Furthermore, objective biomarkers like multiscale entropy (MSE) may offer new ways to measure effects. Bell et al. (2012) reported that homeopathic remedies such as Coffea cruda and Nux vomica produced significant, remedy-specific alterations in the MSE of slow-wave sleep electroencephalograms (EEG) in individuals with coffee-induced insomnia [44]. In addition, a retrospective observational study by Coppola and Montanaro (2013) reported statistically significant reductions in anxiety symptoms, improvements in total sleep time, and a decrease in nocturnal awakenings following the use of a complex homeopathic medicine [45]. While the open-label, retrospective design significantly limits the strength of these conclusions, the findings suggest a clinical correlation between the intervention and improved sleep. These results align with the overall positive trend in our synthesis but also underscore the variability in outcomes and methodological quality.

An earlier systematic review by Cooper and Relton (2010), which included only four RCTs, reported no statistically significant effect of homeopathic medicines for insomnia [14]. A subsequent systematic review by Ernst (2011) included six RCTs and found that homeopathy was not an effective treatment for insomnia [15]. In contrast, our findings align with more recent reviews by Jadhav and Prakash (2023) and Aphale, Sharma, and Shekhar (2024), which found potential benefits of homeopathic interventions for insomnia in adults and children [17, 13]. Nevertheless, our positive effect size stands in contrast to the broader body of literature on complementary and alternative interventions for sleep. In an umbrella review of complementary and alternative interventions for insomnia, Baglioni et al. (2020) reported insufficient evidence to support homeopathy, contrary to other interventions like melatonin and meditative movement therapies [46]. Major consensus guidelines recommend Cognitive Behavioral Therapy for Insomnia (CBT-I) as a highly effective treatment supported by a strong, high-quality evidence base [47]. This discrepancy highlights the uncertainty within the literature on homeopathy for insomnia and the frequent lack of replication for positive findings.

On a broader level, the evidence for the efficacy of homeopathy, in general, is limited, which raises doubts about its clinical effectiveness. In a meta-analysis, Shang et al. (2005) revealed that when analysis was restricted to larger, higher-quality trials across various health conditions, the effects of homeopathy were indistinguishable from those of a placebo [48]. While Shang et al. concluded that the effects of homeopathy are indistinguishable from placebo, a subsequent sensitivity analysis by Lüdtke and Rütten (2008) challenged this finding, demonstrating that when all high-quality trials are considered, homeopathy shows a significant effect beyond placebo [49]. This counter-analysis argues that the definitive negative conclusion of Shang et al. was not robust. Instead, it was highly sensitive to an arbitrary sample size threshold, unduly influenced by a single outlier trial, and undermined by high heterogeneity, which makes firm conclusions from any small subset of trials unreliable.

Likewise, a comprehensive evidence assessment by the National Health and Medical Research Council (NHMRC) of Australia, which evaluated 57 systematic reviews on homeopathy across 68 health conditions, concluded that there is no reliable evidence that homeopathy is effective for treating numerous health conditions [50]. This is in line with other major analyses, including a report by the European Academies’ Science Advisory Council (EASAC) that found no strong, replicable evidence for homeopathy beyond placebo effects [51]. Our meta-analysis is the first one of its kind as none of the earlier systematic reviews mentioned above on the topic conducted a meta-analysis. Furthermore, it yielded a large and statistically significant pooled effect size (SMD = 0.81) favouring homeopathic interventions for sleep improvement. While this result is derived from a robust methodological approach, its interpretation requires careful consideration of the included studies’ characteristics. The magnitude of the effect should be viewed in the context of the identified risk of bias and substantial heterogeneity, which may inflate the estimate. Therefore, the findings are best interpreted not as definitive proof of efficacy, but as a strong signal from the current body of evidence, underscoring the need for more rigorous, high-quality primary studies to confirm these results.

### Limitations of Findings

The large and significant heterogeneity (I² = 86.04%, p < 0.0001) suggests that the studies may not be estimating a single true effect. This inconsistency is a significant limitation, likely explained by differences in homeopathic regimens, such as the use of different and often individualized remedies [52]. The nature of personalized care also presents a scientific dilemma; in studies of individualized homeopathic treatments, it is notoriously difficult to blind participants and providers because creating a convincing placebo is challenging. Furthermore, a high risk of bias is a common shortcoming of trials in complementary and alternative medicine [53]. Additionally, individuals who seek homeopathy may have pre-existing beliefs in its effectiveness, which could greatly enhance placebo effects [54, 55].

Above all, a profound limitation is the high probability of publication bias (Egger test = 0.0079). In contested fields like homeopathy, the ‘file drawer effect’ where small studies with null results remain unpublished is probable and threatens the validity of any synthesis [56]. The asymmetry of the funnel plot strongly suggests that the large overall effect is likely driven by an over-representation of small, positive studies. The pooled SMD is almost certainly an overestimate of the true effect, and it is plausible that if all conducted studies were published, the effect would be markedly smaller and potentially non-significant. When this publication bias is considered alongside the high heterogeneity and risk of bias, the cumulative certainty of the evidence for the pooled effect must be considered low.

### Limitations of the Review Process

Our review process has several limitations that could have influenced the findings.. Firstly, we only included publications in English, which introduces a potential language bias [57]. Secondly, while we searched multiple databases and clinical trial registries, we found that some trials are still in progress, and in some completed trials, the results have not been published [58, 59, 60, 61, 62], which warrants an update of this meta-analysis in near future.

### Implications for Policy and Practice

This review highlights that contextual elements, such as the personalized care and patient-centred approaches typical of homeopathic practice, may produce clinically meaningful improvements for sleep disorders [63, 64, 65]. The results also emphasise the importance of considering homeopathic interventions as an alternative therapeutic option for sleep disorders. The analysis further demonstrates a clear therapeutic effect that affirms empathetic, patient-centred care, of which the individualised homeopathic consultations is a powerful example.

However, the assessment of homeopathic evidence requires a decolonial lens that questions the epistemic hegemony of randomized controlled trials (RCTs) as the sole arbiter of scientific validity [66]. Such an approach recognizes that whole medical systems like homeopathy and other non-Eurocentric modalities are grounded in distinct ontological and epistemological principles that may not align neatly with conventional biomedical research designs [67, 68]. The fundamental homeopathic concept of prescribing a remedy based on a patient’s unique symptoms is incompatible with RCTs, which require standardized interventions for homogeneous groups. Moreover, homeopathic remedies are individualized, meaning the remedy, potency, and dose can be adjusted based on the patient’s response [69]. Blinding can also be compromised, as homeopaths may be able to identify patient reactions to a dose and accurately determine whether they are administering homeopathy or a placebo. These challenges make it difficult to conduct high-quality RCTs. Furthermore, a shortage of research centres and funding for homeopathy limits the number of trained researchers and quality trials. Consequently, policymakers, healthcare administrators, and funders are left without the high-quality evidence needed to settle the decades-long debate on efficacy of homeopathic medicine.

Additionally, regulatory bodies acknowledge this complexity, even if they do not exempt these systems from demonstrating safety and efficacy. For example, the U.S. Food and Drug Administration’s Botanical Drug Development Guidance [70] allows adaptive approaches when prior human use provides supportive evidence, while its Homeopathic Drug Products Guidance [71] applies the same regulatory standards as conventional drugs but exercises enforcement discretion in certain contexts. Similarly, the European Medicines Agency, the UK’s Medicines and Healthcare products Regulatory Agency and Health Canada permit simplified registration pathways for homeopathic products that make no therapeutic claims, relying instead on historical evidence of traditional use (72, 73, 74). The World Health Organization likewise encourages culturally appropriate and epistemologically plural approaches to the evaluation of traditional and complementary medicine (75). Collectively, these frameworks demonstrate a growing global recognition that the assessment of alternative medical systems must account for their unique evidentiary paradigms while maintaining safety and public health standards.

### Implications for Future Research

This review highlights that the crucial need is not simply for more trials, but for methodologically sound trials designed to answer long-standing questions. Future research should be developed to directly address the limitations of the current literature. To address these shortcomings, it is critical to consider the recommendations by Gaertner et al. (2023) on conducting RCTs in homeopathy [69]. These recommendations begin with defining the study goal as either more explanatory (e.g., to test the specific effect of a homeopathic medicine) or more pragmatic (e.g., to evaluate the effectiveness of homeopathic care as a whole system). It is also crucial to consider usual care or standard treatment as a comparator, rather than only a placebo, to test the effect of various homeopathic interventions.

Furthermore, the limitations of existing trial designs may be addressed by employing novel designs, such as an observational study to identify patients who respond to homeopathy, who are then randomized into a double-blind, placebo-controlled crossover trial [68]. It is also recommended to use a pre-defined list of remedies or a computerized algorithm based on major symptom groupings to constrain the number of medicines employed, allowing for easier replication while retaining some degree of individualization. Other recommendations include recruiting a large number of patients, providing homeopathic interventions to a random subset of this cohort, and comparing their outcomes to those who only received usual care. Likewise, multi-arm trials that include homeopathy, placebo, and usual care are another suggested approach [68]. To ensure robust blinding, using a centralized pharmacy to prepare and dispense blinded treatments is a feasible alternative. Finally, it is advised to use double-dummy designs and a combination of disease-specific measures and quality-of-life questionnaires.

### Conclusion

This systematic review and meta-analysis synthesized evidence from 12 studies, including nine randomized controlled trials (RCTs), to evaluate the effectiveness of homeopathic interventions for insomnia. Our meta-analysis of the RCTs identified a statistically significant and large overall effect (SMD = 0.81, 95% CI [0.24, 1.38]), suggesting that homeopathy may help in improving sleep outcomes. The three non-randomized studies also reported statistically significant improvements in sleep quality following homeopathic treatment. However, this result should be interpreted with caution, because the certainty of the evidence was low according to the GRADE assessment, high heterogeneity (I² = 86.04%), and strong evidence of publication bias. To resolve the current uncertainty, there is an urgent need for large, methodologically rigorous, and transparently reported randomized controlled trials and appropriate study designs.

## Supplementary materials

### Appendix I

#### Appendix II: Figures

Figure 1: Preferred Reporting Items for Systematic Reviews and Meta-Analyses (PRISMA) Flow Diagram

Figure 2: Risk of Bias Assessment (Parallel Arm RCTs)

Figure 3: Risk of Bias Assessment Summary (Parallel Arm RCTs)

Figure 4: Risk of Bias Assessment (Crossover Trial)

Figure 5: Risk of Bias Assessment Summary (Crossover Trial)

Figure 6: Risk of Bias Assessment (Observational Studies)

Figure 7: Risk of Bias Assessment Summary (Observational Studies)

Figure 8: Meta-Analysis of Interventions on Sleep Outcomes

Figure 9: Funnel Plot for Publication Bias Assessment

#### Appendix III: Tables

Table 1: Study Characteristics

Table 2: Intervention Characteristics

Table 3: TIDier Checklist for Interventions

Table 4: Summary of the Results of Individual Studies

Table 5: GRADE Summary of findings

## Availability of data, code, and other materials

All meta-analytic data, the master data extraction spreadsheet, completed risk of bias tools, and the analysis scripts (R) are publicly available as supplementary files with this manuscript.

## Supporting information

Search Strategy

PRISMA check list

## Data Availability

https://www.openicpsr.org/openicpsr/project/238782/version/V1/view

## Acknowledgments

We acknowledge the Systematic Reviews Network (SRN) Mentorship Programme 2025 Yasith Mathangasinghe for his supervision, Julia Ribeiro for her mentorship.

Special thanks to Dr. Saima Aleem (Khyber Medical University)

---

Figure 8: Forest Plot of Standardized Mean Differences for Interventions on Sleep Outcomes The plot displays the standardized mean difference (SMD) and 95% confidence interval (CI) for each individual study, as well as the overall pooled effect from a random-effects meta-analysis. The black squares represent the point estimate of the SMD for each study, with their size proportional to the study’s weight in the meta-analysis. The horizontal lines extending from the squares indicate the 95% CI. The diamond at the bottom represents the overall pooled SMD and its 95% CI. A positive SMD indicates a favourable outcome for the intervention.

