## Supplementary material for "Effectiveness of Homeopathic Interventions for Insomnia and Sleep Disorders: A Systematic Review and Meta-Analysis": Figures

**Appendix II: Figures**

Figure 1: Preferred Reporting Items for Systematic Reviews and Meta-Analyses (PRISMA) Flow Diagram

**Identification of studies via databases and registers**

Records removed *before screening*:

Duplicate records removed (n =317)

Records identified from*: (n= 1304)

Databases (Total =1292)

PubMed (n=168)

EuroPMC (n=192)

Cochrane Library (n=141)

EBSCO host (n=136)

Google Scholar (n=573)

DOAJ (n=54)

ProQuest (n=28)

Registers (n =12)

ICTRP (n=8)

ClinicalTrials.Gov (n=4)

**Identification**

Records excluded:

Background article (n =405 )

Wrong population (n =187)

Wrong outcome (n=60)

Wrong drug (n= 72)

Wrong study design (n=119 )

Wrong publication type (n=70)

Wrong study duration

(n= 7)

Records excluded**

(n = 920)

Records screened

(n =987)

Records not retrieved

(n = 0)

Records sought for retrieval

(n =67)

**Screening**

Records excluded (n=56)

Background article (n = 2)

Wrong population (n =4)

Wrong outcome (n = 3

Wrong drug (n= 15)

Wrong study design (n=7)

Wrong publication type (n=19)

Wrong study duration (n=6)

Records assessed for eligibility

(n =67)

**Included**

Studies included in review

(n =12)

Reports of included studies

(n = 0)

**Stu**

Figure 2: Risk of Bias Assessment (Parallel Arm RCTs)

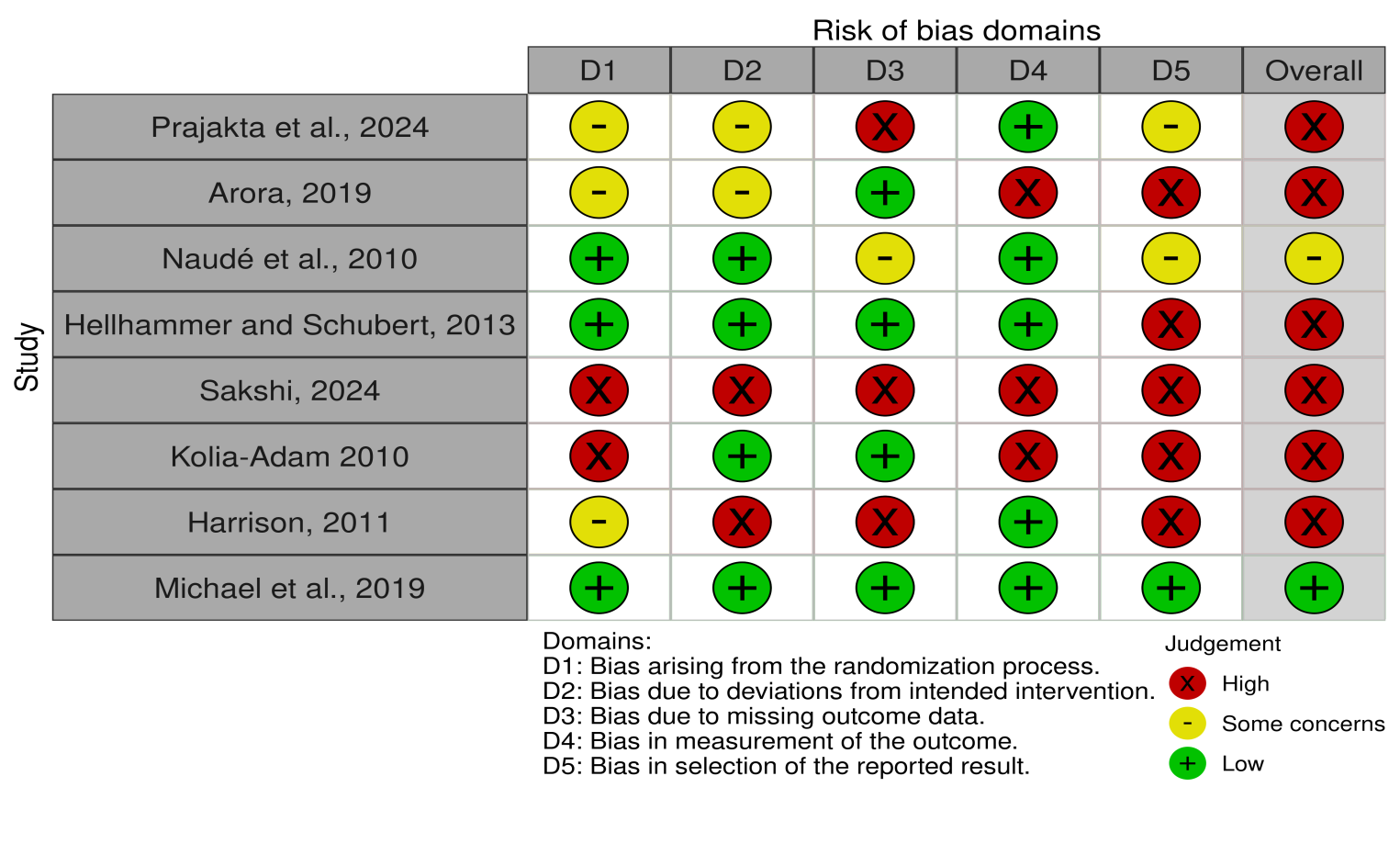

Figure 3: Risk of Bias Assessment Summary (Parallel Arm RCTs)

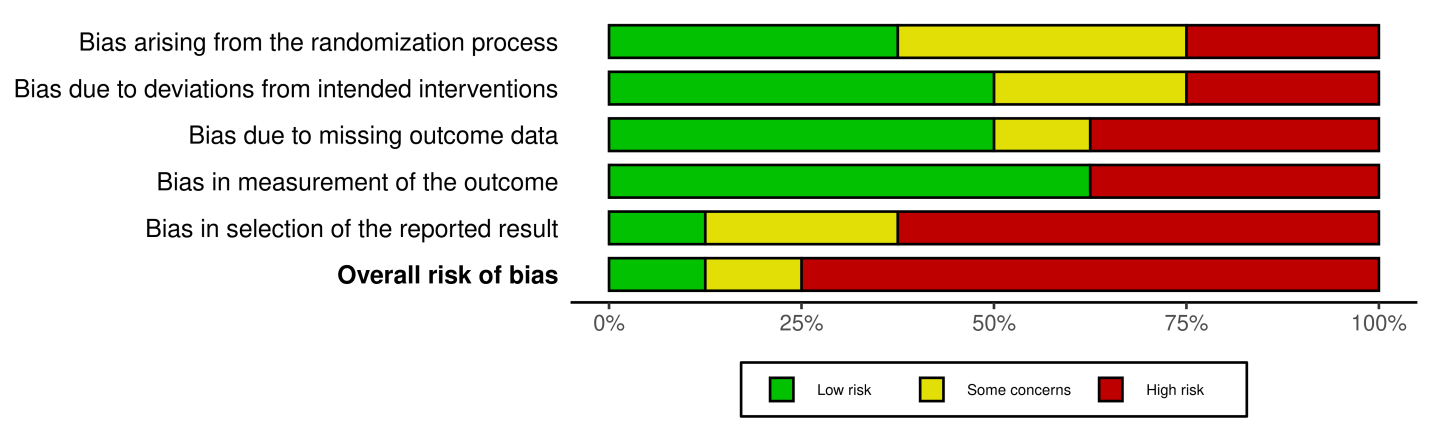

Figure 4: Risk of Bias Assessment (Crossover Trial)

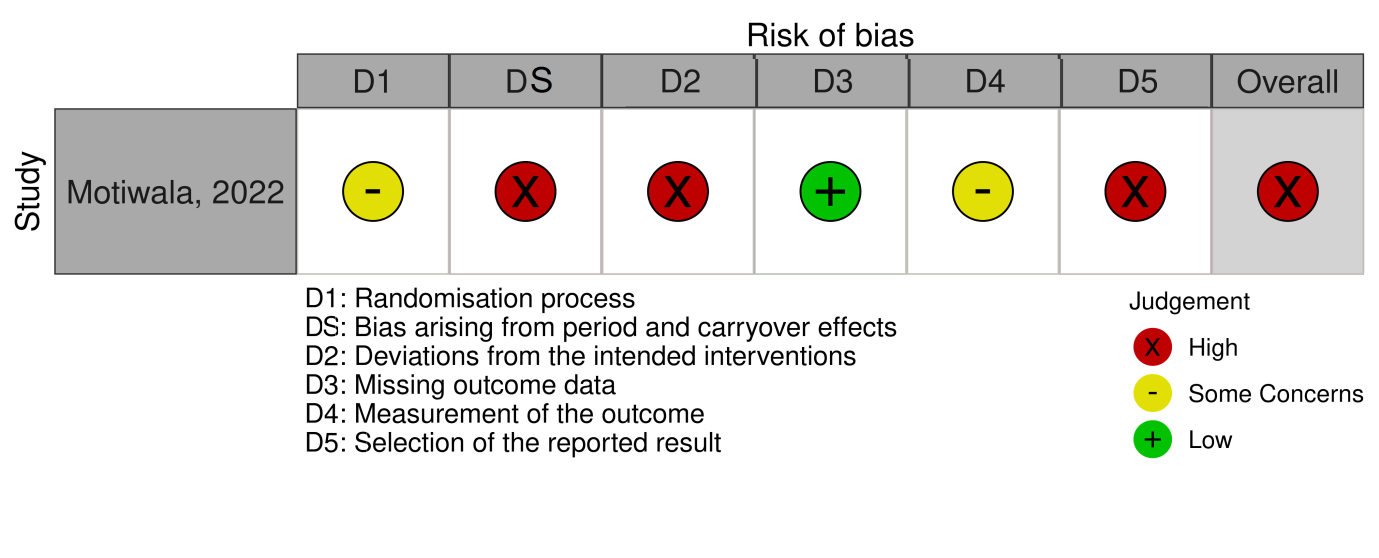

Figure 5: Risk of Bias Assessment Summary (Crossover Trial)

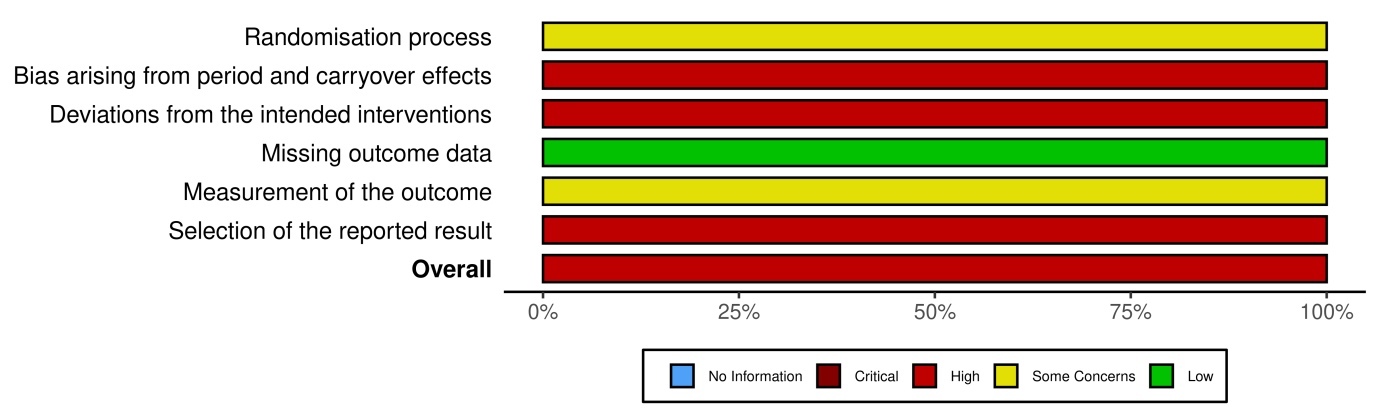

Figure 6: Risk of Bias Assessment (Observational Studies)

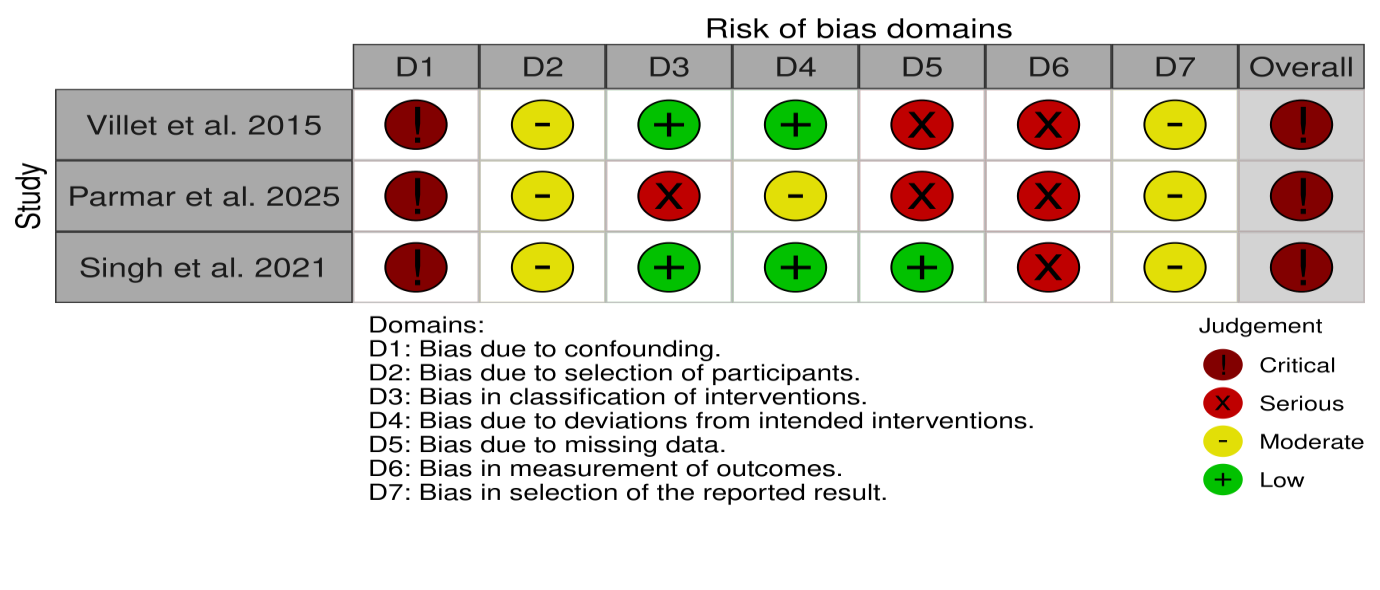

Figure 7: Risk of Bias Assessment Summary (Observational Studies)

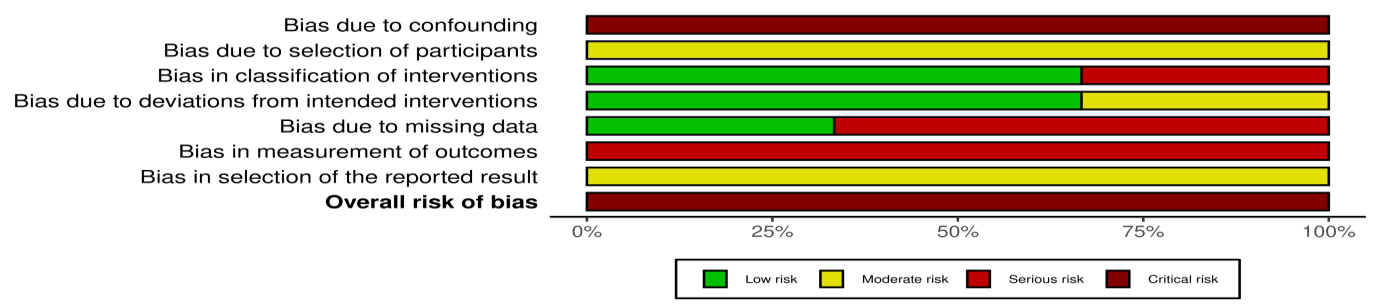

Figure 8: Meta-Analysis of Interventions on Sleep Outcomes

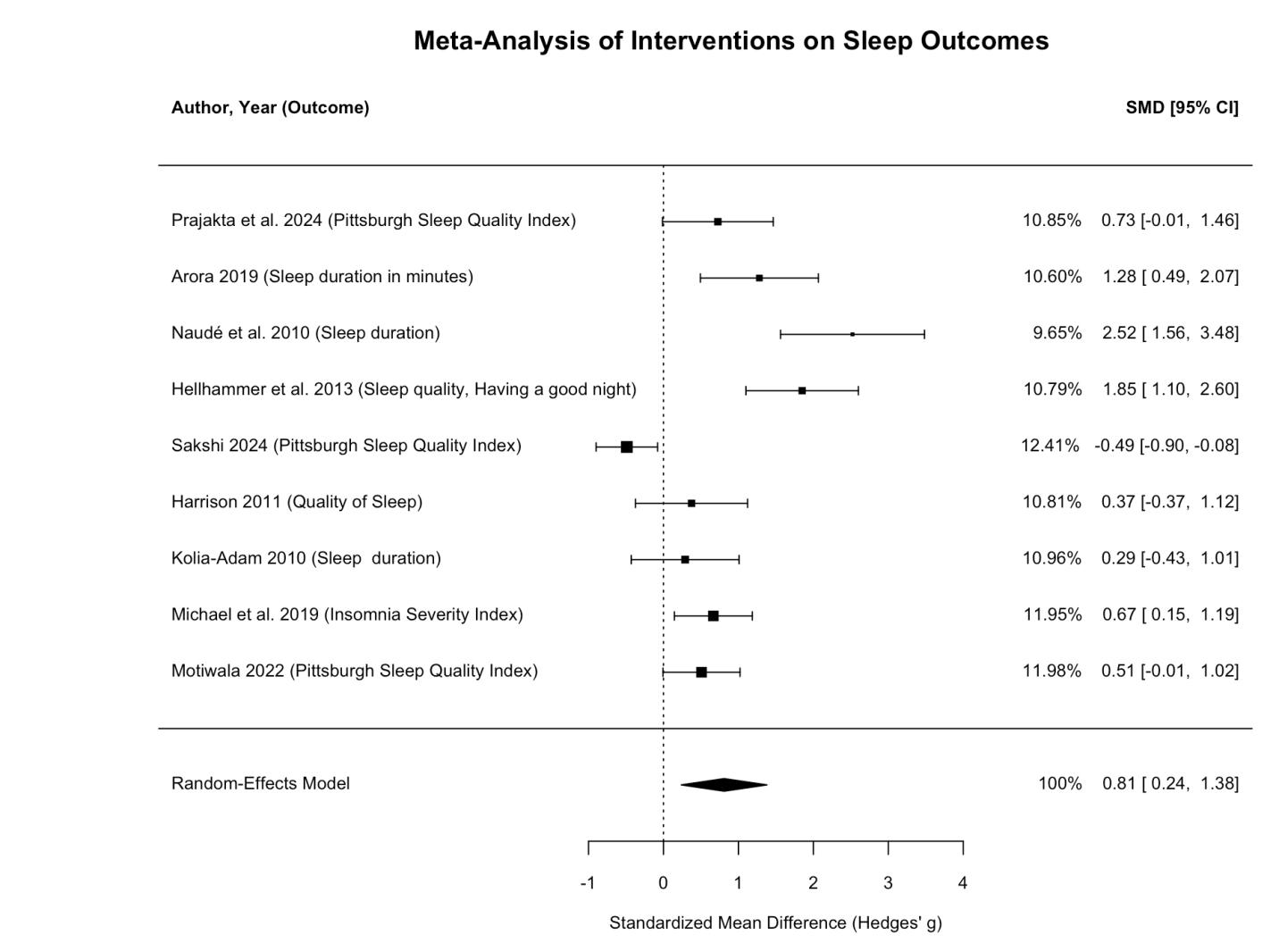

Figure 9: Funnel Plot for Publication Bias Assessment
