## Supplementary material for "Effectiveness of Homeopathic Interventions for Insomnia and Sleep Disorders: A Systematic Review and Meta-Analysis": TABLES

**Appendix II**

**Table 1: Study Characteristics**

| **Author (Year)** | **Country** | **No of Participants** | **Age Range** | **Primary clinical characteristics** | **Secondary clinical characteristics** | **Intervention 1/ Frequency** | **Intervention 2** | **Control** | **Outcome** | **Assessment time** |
| --- | --- | --- | --- | --- | --- | --- | --- | --- | --- | --- |
| Prajakta et al., 2024 | India | 35 | 20-45 years | Primary insomnia | NR | Individualized Homeopathic medicine Remedial Measures (SCT, SRT)/ 6 weeks | NA | Placebo + Remedial Measures | (PSQI) | Baseline, 2 weeks, 4 weeks, 6 weeks |
| Arora, 2019 | India | 30 | 25-45 years | Primary insomnia | NR | Eschscholzia californica MT/ 6 weeks | Passiflora incarnata MT/ 6 weeks | NA | Sleep duration (minutes) | Baseline, day 10, day 20, day 30, day 40 |
| Naudé et al., 2010 | South Africa | 33 | 20-58 years | Chronic primary insomnia (DSM-IV TR 307.42) | NR | Homeopathic simillimum/ 4 weeks | NA | Placebo | Sleep duration (hrs/ week) Sleep Diary, | Baseline, 2 weeks, 4 weeks |
| Hellhammer et al., 2013 | Germany | 40 | 30-50 years | Women regularly experiencing impaired well-being when stressed | Stress-related symptoms: uneasiness, nervousness, attention deficit, tension, fatigue, sleep disorders, headaches, lack of concentration, gastrointestinal disorders | Dysto-loges S (combination homeopathic remedy)/ 2 weeks | Placebo/ 2 Months | Placebo | Sleep quality, Having a good night | Baseline, day 7, day 14, day 15 |
| Villet et al., 2015 | France | 639 | 18-91 years | Anxiety and/or sleep disorders | NR | Passiflora Compose (PC) only/ 4 weeks | Passiflora Compose (PC) + psychotropics/ 4 weeks | Passiflora Compose (PC) + complementary therapies | (JSS) | Baseline, 4 weeks, |
| Sakshi, 2024 | India | 94 | >60 years | Geriatric patients (>60y), elder abused (Group A) or non-abused (Group B), poor sleep quality (PSQI >6) | NR | Homoeopathic Medicines, Elder Abused/ 4 Months | Homoeopathic Medicines, Non-Abused/ 4 months | NA | (PSQI) | Baseline, 2 months, 4 months |
| Parmar et al., 2025 | India | 40 | 18-70 | Insomnia, | DSM-5 criteria | Individualized homoeopathic medicines/ 3 Months | NA | NA | (ISI). | Baseline, 15 days, 30, 45 days, 60, 75, 90 days |
| Singh et al., 2021 | India | 30 | 60-77 | Geriatric insomnia, age ≥60 | NR | Individualized homeopathic medicines/NR | NA | NA | PSQI | NR |
| Harrison, 2011 | South Africa | 28 | 18-40 years | Psychophysiological onset insomnia in males, sleep onset latency >30 min, hyper arousal | Anxiety, intrusive thoughts, restlessness at bedtime | Homoeopathic complex/ 4 weeks | NA | Placebo drops, matched administration | Quality of sleep | Baseline, 2 weeks, 4 weeks |
| Kolia-Adam, 2010 | South Africa | 30 | 18-50 years | Insomnia (difficulty falling asleep, nervous excitability), duration ≤1 year | NR | Coffea cruda 200cH/ 4 weeks | NA | NR | Sleep duration (hours/night sleep diary) | Baseline, 2 weeks, 4 weeks |
| Michael et al., 2019 | India | 60 | 18-60 years | Chronic insomnia, | NR | Individualized Homeopathic Intervention (IHI)/ 3 months | NR | Placebo | (ISI), Sleep diary | Baseline, 12 weeks |
| Motiwala 2022 | India | 30 | 30-60 years | Insomnia (acute/chronic) | NR | Passiflora incarnata Q (MT+)/ 3 months | NA | Placebo | PSQI score | Baseline, 3 months |

**Abbreviations:** SCT, stimulus control therapy; SRT, stimulus restriction therapy; PSQI, Pittsburg sleep quality Index; JSS, Jenkins Sleep Scale; ISI, Insomnia severity index; M, Mean; SD, Standard deviation; DSM, Diagnostic Statistical Manual of Mental Disorders; MT, Mother Tincture; NR, Not reported; NA, Not Applicable

**Table 2: Intervention Characteristics**

| **Author (Year )** | **Study Design** | **Intervention 1 Title** | **Intervention 1 Description** | **Intervention 2 Title** | **Intervention 2 Description** | **Duration of Intervention** |
| --- | --- | --- | --- | --- | --- | --- |
| Prajakta et al., 2024 | RCT | Individualized Homeopathic Medicine such + Remedial Measures (SCT, SRT) | Constitutional homeopathic medicines individualized per case, Arsenicum album, Nux vomica, Kali phosphoricum, Coffea cruda, and Argentum nitricum. Ignatia amara, Silicea, Medorrhinum, Calcarea carbonica, and Natrum muriaticum plus Stimulus Control Therapy and Sleep Restriction Therapy | NA | NA | 6 weeks |
| Arora, 2019 | RCT | Eschscholzia californica MT | 20 drops mother tincture (MT) at bedtime | Passiflora incarnata MT | 20 drops mother tincture (MT) at bedtime | 6 weeks |
| Naudé et al., 2010 | RCT | Homeopathic simillimum | Individualized remedy (various simillima/potencies; 3 single-dose lactose powder sachets per consultation, one taken nightly) | NA | NA | 4 weeks |
| Hellhammer et al., 2013 | RCT | Dysto-loges S (combination homeopathic remedy) | Tablets containing: Passiflora incarnata TM (13mg), Gelsemium D4 (39mg), Reserpinum D6 (31.2mg), Coffea D6 (33.3mg), Veratrum D6 (33.3mg); three tablets daily for 14 days | Placebo | NA | 2 weeks |
| Villet et al., 2015 | observational, longitudinal study | Passiflora Compose (PC) only | PC (oral drops, pillules, tablets): Passiflora incarnata 3DH, Ignatia amara 4CH, Coffea cruda 5CH, Nyckterinia 4CH, Tellurium metallicum 5CH, Phosphoricum acidum 7CH, Palladium metallicum 5CH, Magnesium metallicum 5CH. Dose: most patients 5 granules 2x/day for 1 month. | Passiflora Compose (PC) + psychotropics | PC + at least one psychotropic (anxiolytic/benzodiazepine/hypnotic/antidepressant/neuroleptic) | 4 weeks |
| Sakshi, 2024 | RCT | Homoeopathic Medicines, Elder Abused (Group A) | Individualized homoeopathic medicines based on totality of symptoms (prescribing table available); selection using Synthesis 9.1 repertory, RADAR OPUS software, dose and repetition per patient susceptibility. Palladium, Phosphorus ,Natrum mur. Stramonium, Mercurius sol., Lachesis, Staphysagria, Ignatia, Pulsatilla, Aurum met, Carcinosin, Lycopodium, Bryonia, Opium, Sulphur, Causticum, Baryta carb, Nux vomica, Platina | Homoeopathic Medicines, Non-Abused (Group B) | Individualized remedy Nux vomica, Calcarea carb., Gelsemium, Pulsatilla, Plumbum, Opium, Mercurius solubilis, Coffea, Sepia, Arsenic album, Sulphur, Silicea, Lachesis, Causticum, Kali carbonicum, Bryonia, Hyoscyamus, Aurum met., Phosphorus, Belladonna, Thuja, Tuberculinum, Iodium | 4 months |
| Parmar et al., 2025 | RCT | Individualized homoeopathic medicines | Prescribed as per homoeopathic principles, using Zomeo Elite Software for repertorization, selection and repetition individualized per totality of symptoms; e.g., Nux vomica (17.5%), Pulsatilla (15%), Natrum mur/Cocculus (7.5% each), Sulphur, Coffea cruda, Staphysagria, Phosphorus, Kali phosphoricum (5% each), Lachesis, Gelsemium and Ignatia (2.5%) | NA | NA | 3 months |
| Singh et al., 2021 | Prospective, observational study | Individualized homoeopathic medicines | Medicine chosen per totality of symptoms and homoeopathic principles; Natrum muriaticum , Lachesis , Pulsatilla nigricans , Arsenicum album, Sulphur, Calcarea phosphorica; dosage/potency individualized, procurement from GMP-certified pharmacy | NA | NA | NG |
| Harrison, 2011 | Double-blind, placebo-controlled, matched pairs study | Homoeopathic complex | Ambra grisea 6cH, Arsenicum album 6cH, Coffea cruda 6cH, Delphinium staphisagria 6cH, Ignatia amara 6cH, Lycopodium clavatum 6cH, Passiflora incarnata 6cH, Valeriana officinalis 6cH | NA | NA | 4 weeks |
| Kolia-Adam, 2010 | Double-blind placebo-controlled study | Coffea cruda 200cH | 10 drops of Coffea cruda 200cH (liquid), under tongue at bedtime, daily for 4 weeks. | NR | NR | 4 weeks |
| Michael et al., 2019 | RCT | Individualized Homeopathic Intervention (IHI) | Intervention was planned as administration of indicated homeopathic medicines in centesimal or 50 millesimal potencies and in individualized dosage, as decided appropriate to the case or condition. In centesimal potencies, each dose consisted of 4 cane sugar globules no. 30, moistened with a single drop of the indicated medicine, preserved in 90% v/v ethanol; repetition depending upon the individual requirement of the case and as per homeopathic principles. In 50 millesimal scale, a single medicated cane sugar globule of poppy seed size (no. 10) was dissolved in 90 ml of distilled water with addition of 2 drops of 90% v/v ethanol; 16 doses to be marked on the vial; each dose of 5 ml to be taken after 10 uniformly forceful downward strokes to the vial in 45 ml normal water in a clean cup, to stir well, to take 5 ml of this liquid orally, and to discard rest of the liquid from the cup. Each dose was directed to be taken orally on clean tongue with empty stomach. The most frequently used medicines were Natrum muriaticum(n=10; 43.5%), Nux vomica (n=6, 26.1%), Calcarea carbonicum, Lycopodium clavatum, Mercurius solubilis, Phosphorus, and Sulphur (n=4 each; 17.4%), Pulsatilla pratensis, Sepia succus, and Thuja occidentalis (n=3 each; 13.0%). | NR | NR | 3 months |
| Motiwala 2022 | Placebo controlled cross over trial | Passiflora incarnata Q (Mother tincture) | 30-60 drops of mother tincture orally; lifestyle modifications advised | NA | NA | 3 Months |

**Abbreviations**: RCT, Randomized Controlled Trial; MT, Mother Tincture; SCT, stimulus control therapy; SRT, stimulus restriction therapy, PC, Passiflora compose

**Table 3: TIDier Checklist for Interventions**

| **Intervention Title/ Name** | **Why** | **What** | | **Intervention Provider** | **Mode of Delivery** | **Study Setting** | **Duration** | **Any Modifications** | **Intervention Adherence/ fidelity** |
| --- | --- | --- | --- | --- | --- | --- | --- | --- | --- |
|  |  | **Material Used** | **Procedure** |  |  |  |  |  |  |
| Individualized Homeopathic Medicine such + Remedial Measures (SCT, SRT) | To assess homeopathy's efficacy in treating primary insomnia. | Arsenicum album, Nux vomica, Kali phosphoricum, Coffea cruda, & Argentum nitricum. Ignatia amara, Silicea, Medorrhinum, Calcarea carbonica, & Natrum muriaticum plus Stimulus Control Therapy & Sleep Restriction Therapy | Detailed case histories were obtained following a standardized format. | NG | Oral & face to face | OPD, Dr. D. Y. Patil Homoeopathic Medical College and Research Centre, Pune, India | 6 weeks | Individualized per case, potency selection by protocol included 30C, 200C, and 1M | All participants were advised to adhere to Stimulus Control Therapy and Sleep Restriction Therapy and to attend follow-up assessments every 15 days. |
| Eschscholzia californica MT & Passiflora incarnata MT | To compare the efficacy of E. californica MT and Passiflora incarnata MT in Insomnia. | Dosage 20 drops of both homeopathic medicines were given bed time…. ( | Eschscholzia californica MT - 16 Subjects Passiflora incarnata MT – 14 Subjects. Patient were screened and enrolled. | NG | Oral | OPD, Bharati Vidyapeeth University; Institute of Management and Research, New Delhi, India | 6 weeks | NR | NR |
| Homeopathic simillimum | To evaluate the efficacy of homeopathic simillimum in the treatment of chronic primary insomnia. | Individualized remedy (various simillima/potencies; 3 single-dose lactose powder sachets per consultation, one taken nightly) | At the first consultation, a full homeopathic case history was taken and a physical examination was performed | An independent dispenser at the DUT Homeopathic Day Clinic | Oral (sublingual lactose powder sachets) | Durban University of Technology Homeopathic Day Clinic, South Africa | 4 weeks | Potency individualized per participant 30CH, 200CH, 1M, 10M per protocol | NR |
| Dysto-loges S (combination homeopathic remedy) | To investigate stress dampening effects of the homeopathic combination remedy dysto-loges-S on physiological and psychological measures during acute stress; to assess effects on sleep and quality of life. | Tablets containing: Passiflora incarnata TM (13mg), Gelsemium D4 (39mg), Reserpinum D6 (31.2mg), Coffea D6 (33.3mg), Veratrum D6 (33.3mg); three tablets daily for 14 days | All participants were asked to take three tablets of dysto-loges S or the matching placebo daily for the duration of 14 days | Daarco contract research organization | Oral (dissolved in mouth, 1 before each meal) | Daacro, Trier, Germany | 2 weeks | None | NR |
| Passiflora Compose (PC) only,  &  Passiflora Compose (PC) + psychotropics | To describe the socio-demographic characteristics and clinical progression of patients prescribed homeopathic medicine Passiflora Compose (PC) for anxiety and/or sleep disorders (SDS) | PC (oral drops, pillules, tablets): Passiflora incarnata 3DH, Ignatia amara 4CH……. | At the inclusion visit, GPs completed an inclusion form recording data on: the socio-demographic characteristics of the patients; the reason(s) for consultation | General Practitioners | Oral, self-administered | General practitioners’ offices (France, 98 GPs) | 4 weeks | NR | 4 weeks after inclusion, patients were assessed using HAM-A scale and follow-up form questions. |
| Homoeopathic Medicines, Elder Abused (Group A) & Non Abused (Group B) | To assess the role of homoeopathic medicines in improving sleep quality in elder abused patients as compared to non-abused geriatric patients. | Individualized homoeopathic medicines based on totality of symptoms (prescribing table available) | Patients were selected in two groups: Group A (Elder abused), group B (Non abused) geriatric patients with reduced sleep quality. . | NG | Oral | Bikaner OPD of MNHMC & RI | 4 Months | NR | NR |
| Individualized homoeopathic medicines | To assess the role of homoeopathic medicines in treating cases of insomnia. | Prescribed as per homoeopathic principles, using Zomeo Elite Software for repertorization, selection and repetition individualized per totality of symptoms……… | Final selection of remedy is done upon the basis of totality of symptoms by consulting with Homoeopathic | OPD Jawaharlal Nehru Homoeopathic Medical College and Hospital | Oral | OPD Jawaharlal Nehru Homoeopathic Medical College and Hospital | 3 Months | NR | Severity of symptoms of insomnia was assessed with ISI at baseline and at every follow ups. Follow up was taken at interval of 15 days |
| Individualized homeopathic medicines | To evaluate the effectiveness of homeopathic medicine in sleep disorders in geriatric age group | Medicine chosen per totality of symptoms and homoeopathic principles; Natrum muriaticum , Lachesis , Pulsatilla nigricans , Arsenicum album, Sulphur, Calcarea phosphorica; | Homoeopathic Medicine was selected per Homoeopathic principles given in edition of Organon of Medicine….. | Pharmaceutical company | Oral | OPD of Dr. Girendra Pal Homoeopathic Hospital and Research Centre, Jaipur | Not explicitly stated (study duration 6 months, not all patients duration given) | Medicine and/or potency changed as per patient response and principles | NG |
| Homoeopathic complex | To determine the efficacy of a homoeopathic complex on psychophysiological onset insomnia in male participants | Ambra grisea 6cH, Arsenicum album 6cH, Coffea cruda 6cH, Delphinium staphisagria 6cH, Ignatia amara 6cH, Lycopodium clavatum 6cH, Passiflora incarnata 6cH, Valeriana officinalis 6cH | During first visit, the researcher explained the requirement and details of the study | Homeopathy Health Clinic | Oral (5 drops sublingual, twice evening) | University of Johannesburg, Doornfontein campus clinic and local advertisements | 4 weeks | NR | Follow-up consultation occurred at week 2 and week 4 |
| Coffea cruda 200cH | To determine the efficacy of Coffea cruda 200cH in the treatment of insomnia characterized by difficulty in falling asleep; | 10 drops of Coffea cruda 200cH (liquid), under tongue at bedtime, daily for 4 weeks. | Participants received a 50 ml bottle of medications in liquid | Homeopathy Health Clinic | Oral, sublingual drops | University of Johannesburg Health Clinic and community (advertisements) | 4 weeks | NR | Sleep diaries were checked to promote compliance and case histories were taken at week 2 and week 4. |
| Individualized Homeopathic Intervention (IHI) | The primary objective of this study was to evaluate the efficacy of IHIs (individualized homeopathic interventions) as compared with placebo in the treatment of primary insomnia. | Intervention was planned as administration of indicated homeopathic medicines in centesimal or 50 millesimal potencies and in individualized dosage…. | Medicines were obtained from manufacturing Practice certified firms | Qualified homeopathic physicians. | Oral (liquid/globules) | OPD of National Institute of Homoeopathy (NIH). Kolkata, West Bengal, India. | 3 months | NR | NG |
| Passiflora incarnata Q (Mother tincture) | Evaluate efficacy of Passiflora incarnata Q vs. placebo in insomnia | Intervention with homoeopathic medicine, Passiflora incarnata mother tincture. lifestyle modifications advised. | Known patients of insomnia identified through online and offline surveys. | NR | Oral (liquid) | OPD/IPD of college, health camps, private practice, online survey | 3 months | NR | NR |

**Table 4: Summary of the Results of Individual Studies**

| **Study ID** | **Outcome** | **Timings of Outcome** | **Intervention Mean** | **Intervention SD** | **Comparator Mean** | **Comparator SD** |
| --- | --- | --- | --- | --- | --- | --- |
| Prajakta et al., 2024 | Pittsburgh Sleep Quality Index (PSQI) | Week 6 | 14.2 | 2.54 | 16.88 | 4.4 |
| Arora, 2019 | Sleep duration in minutes | Day 40 | 6.63 | 0.29 | 6.04 | 0.58 |
| Naudé et al., 2010 | Sleep duration (hours/week) | Week 4 | 41 | 3.74 | 34 | 1.22 |
| Hellhammer et al., 2013 | Sleep quality, Having a good night (VIS) (mm) | Day 15 | 72.71 | 7.07 | 58.56 | 7.87 |
| Sakshi, 2024 | Pittsburgh Sleep Quality Index (PSQI) | 4 months | 9.26 | 2.23 | 7.98 | 2.92 |
| Harrison, 2011 | Quality of Sleep | 4 weeks | 1.64 | 0.7 | 1.93 | 0.8 |
| Kolia-Adam, 2010 | Sleep duration (hours/night sleep diary) | 4 weeks | 6.59 | 0.72 | 6.37 | 0.76 |
| Michael et al., 2019 | Insomnia Severity Index (ISI) | 3 months | 13.9 | 4.6 | 16.6 | 3.3 |
| Motiwala, 2022 | Pittsburgh Sleep Quality Index (PSQI) | 3 months | 13.23 | 3.21 | 14.9 | 3.28 |
| Villet et al., 2015 | Jenkins Sleep Scale (JSS) | 4 weeks | 10.76 | 4.67 | Not applicable | Not applicable |
| Parmar et al., 2025 | Insomnia Severity Index (ISI) | 3 months | 6.55 | 2.26 | Not applicable | Not applicable |
| Singh et al., 2021 | Pittsburgh Sleep Quality Index (PSQI) | Post-treatment | 6.66 | 2.33 | Not applicable | Not applicable |

| **Table 5: GRADE Summary of findings:** | | | | | | |
| --- | --- | --- | --- | --- | --- | --- |
| **Homeopathic Interventions for insomnia and sleep disorders** | | | | | | |
| **Patient or population:** insomnia and sleep disorders  **Setting:**  **Intervention:** Homeopathy  **Comparison: Placebo, Homeopathy, None** | | | | | | |
| Outcomes | **Anticipated absolute effects^*^** (95% CI) | | Relative effect (95% CI) | № of participants (studies) | Certainty of the evidence (GRADE) | Comments |
|  | **Risk with Placebo** | **Risk with Homeopathy** |  |  |  |  |
| Sleep Quality ( Visual Analogue Scale (VIS)) Scale from: 0 to 100 follow-up: mean 2 weeks | - | SMD **1.85 SD higher** (1.1 higher to 2.6 higher) | - | 40 (1 RCT) | ⨁⨁◯◯ Low^36,a,b^ | Homeopathy may result in a large increase in Sleep Quality. |
| Sleep Duration per week follow-up: mean 40 days | - | SMD **1.28 SD higher** (0.49 higher to 2.07 higher) | - | 30 (1 RCT) | ⨁⨁◯◯ Low^32,b^ | Homeopathy may result in a large increase in Sleep Duration.^b^ |
| Sleep Duration  Scale from: 34 to 41 follow-up: mean 4 weeks | - | SMD **2.52 SD higher** (1.56 higher to 3.48 higher) | - | 30 (1 RCT) | ⨁⨁⨁◯ Moderate^37,b^ | Homeopathy likely results in a large increase in Sleep Duration . |
| Pittsburgh Sleep Quality Index (PSQI) follow-up: mean 16 weeks | - | SMD **0.81 SD higher** (0.24 higher to 1.38 higher) | - | 184 (3 RCTs) | ⨁⨁⨁◯ Moderate^33.35.39,b^ | Homeopathy likely results in a large increase in Sleep Quality. |
| Sleep Duration per night follow-up: mean 4 weeks | - | SMD **0.29 SD higher** (0.43 lower to 1.01 higher) | - | 30 (1 RCT) | ⨁⨁◯◯ Low^31,c^ | Homeopathy may result in a large increase in sleep Duration per night. |
| Quality of Sleep follow-up: mean 4 weeks | - | SMD **0.37 SD higher** (0.37 lower to 1.12 higher) | - | 28 (1 RCT) | ⨁◯◯◯ Very low^34,b,c^ | The evidence is very uncertain about the effect of Homeopathy on quality of Sleep. |
| Sleep Quality Insomnia Severity Index (ISI) follow-up: median 12 weeks | - | SMD **0.67 SD higher** (0.15 higher to 1.19 higher) | - | 60 (1 RCT) | ⨁⨁⨁◯ Moderate^38^,^,b^ | Homeopathy likely results in a large increase in Sleep Quality |
| Sleep Quality Jenkins Sleep Scale (JSS) follow-up: mean 4 weeks |  | mean **10.76 higher** (0 to 0 ) | - | 401 (1 non-randomised study) | ⨁◯◯◯ Very low^40,d^ | The evidence is very uncertain about the effect of Homeopathy on Jenkins Sleep Scale . |
| Sleep Quality Insomnia Severity Index (ISI) follow-up: mean 12 weeks |  | mean **6.55 higher** (0 to 0 ) | - | 40 (1 non-randomised study) | ⨁◯◯◯ Very low^41,e^ | The evidence is very uncertain about the effect of Homeopathy on Sleep Quality |
| Pittsburgh Sleep Quality Index (PSQI) |  | mean **6.66 higher** (0 to 0 ) | - | 30 (1 non-randomised study) | ⨁◯◯◯ Very low^42,f^ | The evidence is very uncertain about the effect of Homeopathy on Sleep Quality |
| ***The risk in the intervention group** (and its 95% confidence interval) is based on the assumed risk in the comparison group and the **relative effect** of the intervention (and its 95% CI).  **CI:** confidence interval; **SMD:** standardised mean difference | | | | | | |
| **GRADE Working Group grades of evidence** **High certainty:** we are very confident that the true effect lies close to that of the estimate of the effect. **Moderate certainty:** we are moderately confident in the effect estimate: the true effect is likely to be close to the estimate of the effect, but there is a possibility that it is substantially different. **Low certainty:** our confidence in the effect estimate is limited: the true effect may be substantially different from the estimate of the effect. **Very low certainty:** we have very little confidence in the effect estimate: the true effect is likely to be substantially different from the estimate of effect. | | | | | | |

#### Explanations

a. High Risk of Bias in Selective Reporting Domain

b. The Egger's regression test for funnel plot asymmetry indicated a statistically significant presence of publication bias (t = 3.68, df = 7, p = 0.0079).

c. High risk of bias in multiple domains

d. The overall risk of bias is critical. The study's design as a single-arm, open-label trial (Domain 1 and 6) is fundamentally flawed and incapable of demonstrating efficacy. The observed improvements are inextricably confounded with natural history, regression to the mean, and placebo/expectation effects.

e. The overall risk of bias is critical, driven primarily by the complete lack of a control group (Domain 1) and the unblinded assessment of a subjective outcome (Domain 6). These flaws create a strong upward bias, making it impossible to conclude that the observed improvement was due to the homeopathic intervention rather than confounding factors.

f. The study is at a critical risk of bias, driven by the single-arm design which makes it impossible to attribute any observed changes to the intervention (Domain 1). This is compounded by a serious risk of bias in the measurement of the unblinded, subjective outcome (Domain 6)

References

31.Kolia-Adam, Naseeha. The efficacy of Coffea Cruda 200cH on insomnia.Unpublished Dissertation; 2010

32..Arora, Dr Anmol. A study to compare the efficacy of Eschscholzia californica MT and Passiflora incarnata MT in insomnia.International Journal of Homoeopathic Sciences; 2019.

33.Sakshi, Dr,Mehrotra. THE ROLE OF HOMOEOPATHIC MEDICINES FOR IMPROVING SLEEP QUALITY IN ELDER ABUSED AND NON-ABUSED GERIATRIC PATIENTS.World Journal of Pharmaceutical Research; 2024

34.Harrison, Caroline Christel, Solomon, Elizabeth Margaret, Pellow, Janice. The effect of a homeopathic complex on psychophysiological onset insomnia in males: a randomized pilot study.Alternative Therapies in Health and Medicine; 2013.

35. Prajakta.et al A Clinical Study on The Efficacy of Homoeopathic Medicines in The Treatment of Primary Insomnia – A Pilot Study”.African Journal of Biomedical Research; 2024.

36. Hellhammer, Juliane, Schubert, Melanie. Effects of a homeopathic combination remedy on the acute stress response, well-being, and sleep: a double-blind, randomized clinical trial.Journal of Alternative and Complementary Medicine (New York, N.Y.); 2013-02.

37.Naudé, David Francis, Stephanie Couchman, Ingrid Marcelline, Maharaj, Ashnie. Chronic primary insomnia: Efficacy of homeopathic simillimum.Homeopathy; 2010-01-01.

38.Michael, James, Singh, Subhas, Sadhukhan, Satarupa, Nath, Arunava, Kundu, Nivedita, Magotra, Nitin, Dutta, Susmit, Parewa, Maneet, Koley, Munmun, Saha, Subhranil. Efficacy of individualized homeopathic treatment of insomnia: Double-blind, randomized, placebo-controlled clinical trial.Complementary Therapies in Medicine; 2019-04-01.

39. Motiwala, ,Pooja. To Study The Efficacy Of Passiflora Incarnata Mother Tincture In Cases of Insomnia, A Placebo Controlled Cross Over Trial.Materia Novum - The Journal of Homoeopathy; 2022.

40.Villet, Stéphanie, Vacher, Véronique, Colas, Aurélie, Danno, Karine, Masson, Jean-Louis, Marijnen, Philippe, Bordet, Marie-France. Open-label observational study of the homeopathic medicine Passiflora Compose for anxiety and sleep disorders.Homeopathy: The Journal of the Faculty of Homeopathy; 2016-02.

41.Parmar, Dr Hiren D., Desai, Dr Poorav, Desai, Dr Kirtida. Sound sleep with sweet pills: A prospective intervational study of homoeopathy in treating insomnia.International Journal of Homoeopathic Sciences; 2025.

42.Singh, Atul. STUDY TO EVALUATE THE EFFECTIVENESS OF HOMEOPATHIC MEDICINE IN SLEEP DISORDERS IN GERIATRIC AGE GROUP.International Journal of AYUSH; 2021.
