## Supplementary material for "Effectiveness of Homeopathic Interventions for Insomnia and Sleep Disorders: A Systematic Review and Meta-Analysis": Search Strategy

PubMed

(("insomnia"[MeSH] OR "sleep initiation and maintenance disorders"[MeSH] OR "insomnia"[Title/Abstract] OR "sleep disorder*"[Title/Abstract] OR "sleep disturbance*"[Title/Abstract])

AND

("homeopathy"[MeSH] OR "homeopathic"[Title/Abstract] OR "homeopath*"[Title/Abstract] OR "valeriana officinalis"[Title/Abstract] OR "passiflora incarnata"[Title/Abstract] OR "coffea cruda"[Title/Abstract] OR "nux vomica"[Title/Abstract] OR "kalium phosphoricum"[Title/Abstract]))

Results: 157

Dates searched: Feb- Mar 2025

Euro PMC

TITLE_ABS:(( sleep* OR insomnia* OR "sleep disturb*" OR "sleep depriv*" OR "sleep disorder*" OR "poor sleep" OR "sleep problem*" "wakefulness" ) AND ( homeopath* OR "homeopathic remed*" OR "homeopathic medicin*" OR "valerian*" OR "passiflora*" OR "kali* phosphoric*" OR "coffea*" OR "nux vomica" OR "aconitum*" OR "arsenicum*" OR "ignatia" OR "lycopodium" OR "stramonium" ) )

RESULTS: 192

Dates Searched: Feb-Mar 2025

Cochrane

Cochrane Library

Population: Insomnia

#1 MeSH descriptor: [Sleep Initiation and Maintenance Disorders] explode all trees

#2 (insomnia or "sleep NEXT initiat" or "sleep NEXT maintain" or "sleep NEXT disorder" or "sleep NEXT disturbance")

#3 #1 or #2

Intervention: Homeopathy

#4 MeSH descriptor: [Homeopathy] explode all trees

#5 (homeopath* or homoeopath*):ti,ab,kw

#6 ("NEXT valerian" or " NEXT passiflor" or "coffea cruda" or "nux vomica" or "ignatia" or "arsenicum album" or "aconitum" or "pulsatilla" or "lycopodium"):ti,ab,kw

#7 #4 or #5 or #6

#8 #3 AND #7

Results: 141

EBSCOhost databases (CINAHL, MEDLINE, APA PsycInfo)

(TI ( insomnia OR "sleep disturbance*" OR "sleep deprivation" OR "sleep disorder*" OR "poor sleep" )

OR	AB ( insomnia OR "sleep disturbance*" OR "sleep deprivation" OR "sleep disorder*" OR "poor sleep" )

OR	SU ( "Sleep Disorders" OR Insomnia ))

AND

( TI ( homeopath* OR "homeopathic remed*" OR "homeopathic treatment" OR "homeopathic medicine*" OR "valeriana officinalis" OR "passiflora incarnata" OR "kali phosphoricum" OR "coffea cruda" OR "nux vomica" ) OR

AB (homeopath* OR "homeopathic remed*" OR "homeopathic treatment" OR "homeopathic medicine*" OR "valeriana officinalis" OR "passiflora incarnata" OR "kali phosphoricum" OR "coffea cruda" OR "nux vomica" ) OR SU ( Homeopathy ))

AND( TI ( treatment OR therapy OR management OR intervention OR trial OR study )

OR	AB ( treatment OR therapy OR management OR intervention )

OR	SU ( "Treatment Outcomes" OR Management OR Therapy ))

AND

(TI ( "sleep quality" OR "sleep duration" OR "sleep latency" OR "sleep improv*" OR "falling asleep" ) OR AB ( "sleep quality" OR "sleep duration" OR "sleep latency" OR "sleep improv*" OR "falling asleep" )

Result:135

Embase

('insomnia'/exp OR 'insomnia':ti,ab OR 'sleep disorder*':ti,ab OR 'sleep disturbance*':ti,ab)

AND

('homeopathy'/exp OR 'homeopathic':ti,ab OR 'homeopath*':ti,ab OR 'valeriana officinalis':ti,ab OR 'passiflora incarnata':ti,ab OR 'coffea cruda':ti,ab OR 'nux vomica':ti,ab OR 'kalium phosphoricum':ti,ab)

Google Scholar

("homeopathy" OR "homeopathic treatment")

AND

("insomnia" OR "sleep disorder")

AND

("sleep latency" OR "wake after sleep onset" ")

Results:547

Directory of Open Access Journals (DOAJ)

title:(homeopath* OR "homeopathic treatment") AND (insomnia OR sleep)

Results: 34

ClinicalTrials.gov

homeopathy AND insomnia:

homeopathy AND sleep

Result: 4

International Clinical Trials Registry Platform (ICTRP)

homeopathy AND insomnia:

homeopathy AND sleep

Results: 8
