## Supplementary material for "Effectiveness of Homeopathic Interventions for Insomnia and Sleep Disorders: A Systematic Review and Meta-Analysis": PRISMA check list

| **Section and Topic** | **Item #** | **Checklist item** | **Location where item is reported** |
| --- | --- | --- | --- |
| **TITLE** | | |  |
| Title | 1 | Identify the report as a systematic review. | Title Page, Line 1: "Effectiveness of Homeopathic Interventions for Insomnia and Sleep Disorders: A Systematic Review and Meta-Analysis" |
| **ABSTRACT** | | |  |
| Abstract | 2 | See the PRISMA 2020 for Abstracts checklist. | Page 2, Abstract |
| **INTRODUCTION** | | |  |
| Rationale | 3 | Describe the rationale for the review in the context of existing knowledge. | Page 4 |
| Objectives | 4 | Provide an explicit statement of the objective(s) or question(s) the review addresses. | Page 4, Final Paragraph |
| **METHODS** | | |  |
| Eligibility criteria | 5 | Specify the inclusion and exclusion criteria for the review and how studies were grouped for the syntheses. | Page 5, "Eligibility Criteria |
| Information sources | 6 | Specify all databases, registers, websites, organisations, reference lists and other sources searched or consulted to identify studies. Specify the date when each source was last searched or consulted. | Page 6, "Information Sources" & Page 8, "Study Selection" (for dates) |
| Search strategy | 7 | Present the full search strategies for all databases, registers and websites, including any filters and limits used. | Page 6, "Information Sources" & Appendix I |
| Selection process | 8 | Specify the methods used to decide whether a study met the inclusion criteria of the review, including how many reviewers screened each record and each report retrieved, whether they worked independently, and if applicable, details of automation tools used in the process. | Page 7, "Selection Process" |
| Data collection process | 9 | Specify the methods used to collect data from reports, including how many reviewers collected data from each report, whether they worked independently, any processes for obtaining or confirming data from study investigators, and if applicable, details of automation tools used in the process. | Page 7, "Data Collection Process" |
| Data items | 10a | List and define all outcomes for which data were sought. Specify whether all results that were compatible with each outcome domain in each study were sought (e.g. for all measures, time points, analyses), and if not, the methods used to decide which results to collect. | Page 6, "Eligibility Criteria (O)" & Page 7, "Data Collection Process" |
|  | 10b | List and define all other variables for which data were sought (e.g. participant and intervention characteristics, funding sources). Describe any assumptions made about any missing or unclear information. | Page 7, "Data Collection Process" |
| Study risk of bias assessment | 11 | Specify the methods used to assess risk of bias in the included studies, including details of the tool(s) used, how many reviewers assessed each study and whether they worked independently, and if applicable, details of automation tools used in the process. | Page 7-8, "Risk of Bias Assessment" |
| Effect measures | 12 | Specify for each outcome the effect measure(s) (e.g. risk ratio, mean difference) used in the synthesis or presentation of results. | Page 9, "Effect Measures" "Effect Size Calculation" |
| Synthesis methods | 13a | Describe the processes used to decide which studies were eligible for each synthesis (e.g. tabulating the study intervention characteristics and comparing against the planned groups for each synthesis (item #5)). | Page 9, "Synthesis Methods" |
|  | 13b | Describe any methods required to prepare the data for presentation or synthesis, such as handling of missing summary statistics, or data conversions. | Page 9, "Effect Size Calculation" |
|  | 13c | Describe any methods used to tabulate or visually display results of individual studies and syntheses. | Page 9, "Synthesis Methods" & Page 10, "Results of Individual Studies" & Appendix II |
|  | 13d | Describe any methods used to synthesize results and provide a rationale for the choice(s). If meta-analysis was performed, describe the model(s), method(s) to identify the presence and extent of statistical heterogeneity, and software package(s) used. | Page 10, "Synthesis Methods" & "Meta-Analysis" |
|  | 13e | Describe any methods used to explore possible causes of heterogeneity among study results (e.g. subgroup analysis, meta-regression). | Page 10, |
|  | 13f | Describe any sensitivity analyses conducted to assess robustness of the synthesized results. | Not Applicable |
| Reporting bias assessment | 14 | Describe any methods used to assess risk of bias due to missing results in a synthesis (arising from reporting biases). | Page 8, "Reporting Bias Assessment" & Page 10, "Heterogeneity and Publication Bias Assessment" |
| Certainty assessment | 15 | Describe any methods used to assess certainty (or confidence) in the body of evidence for an outcome. | Page 11, "Certainty Assessment" |
| **RESULTS** | | |  |
| Study selection | 16a | Describe the results of the search and selection process, from the number of records identified in the search to the number of studies included in the review, ideally using a flow diagram. | Page 11, "Study Selection" & Appendix II, Figure 1 |
|  | 16b | Cite studies that might appear to meet the inclusion criteria, but which were excluded, and explain why they were excluded. | Page 11, "Study Selection" |
| Study characteristics | 17 | Cite each included study and present its characteristics. | Page 11-12, "Study Characteristics" & Appendix III, Table 1 |
| Risk of bias in studies | 18 | Present assessments of risk of bias for each included study. | Page 13-14 "Risk of Bias" & Appendix II, Figures 2-7 |
| Results of individual studies | 19 | For all outcomes, present, for each study: (a) summary statistics for each group (where appropriate) and (b) an effect estimate and its precision (e.g. confidence/credible interval), ideally using structured tables or plots. | Page 15, "Results of Individual Studies" & Appendix III, Table 4 & Appendix II, Figure 8 |
| Results of syntheses | 20a | For each synthesis, briefly summarise the characteristics and risk of bias among contributing studies. | Page 13-14 "Risk of Bias" & Appendix II, Figures 2-7 Page 15, "Results of Syntheses" (Narrative) |
|  | 20b | Present results of all statistical syntheses conducted. If meta-analysis was done, present for each the summary estimate and its precision (e.g. confidence/credible interval) and measures of statistical heterogeneity. If comparing groups, describe the direction of the effect. | Page 17, "Overall Meta-Analysis" & Appendix II, Figure 8 |
|  | 20c | Present results of all investigations of possible causes of heterogeneity among study results. | Page 17, "Overall Meta-Analysis": "The analysis revealed substantial and statistically significant heterogeneity across the studies (Q(df = 8) = 56.72, p < 0.0001; I² = 86.04%)." |
|  | 20d | Present results of all sensitivity analyses conducted to assess the robustness of the synthesized results. | Not conducted |
| Reporting biases | 21 | Present assessments of risk of bias due to missing results (arising from reporting biases) for each synthesis assessed. | Page 17, "Publication Bias" & Appendix II, Figure 9 |
| Certainty of evidence | 22 | Present assessments of certainty (or confidence) in the body of evidence for each outcome assessed. | Page 18, "Certainty of Evidence" & Appendix III, Table 5 |
| **DISCUSSION** | | |  |
| Discussion | 23a | Provide a general interpretation of the results in the context of other evidence. | Page 18-19, "Discussion" |
|  | 23b | Discuss any limitations of the evidence included in the review. | Page 20, "Limitations of Findings" |
|  | 23c | Discuss any limitations of the review processes used. | Page 21, "Limitations of the Review Process" |
|  | 23d | Discuss implications of the results for practice, policy, and future research. | Page 21, "Implications for Policy and Practice" & Page 22 "Implications for Future Research" |
| **OTHER INFORMATION** | | |  |
| Registration and protocol | 24a | Provide registration information for the review, including register name and registration number, or state that the review was not registered. | Page 2, "Registration and Protocol" |
|  | 24b | Indicate where the review protocol can be accessed, or state that a protocol was not prepared. | Page 5, "Registration and Protocol" |
|  | 24c | Describe and explain any amendments to information provided at registration or in the protocol. | Page 5, "Registration and Protocol" |
| Support | 25 | Describe sources of financial or non-financial support for the review, and the role of the funders or sponsors in the review. | Page 1, "Acknowledgments" |
| Competing interests | 26 | Declare any competing interests of review authors. | Page 24, "Competing interests" |
| Availability of data, code and other materials | 27 | Report which of the following are publicly available and where they can be found: template data collection forms; data extracted from included studies; data used for all analyses; analytic code; any other materials used in the review. | Page 23, "Availability of data, code, and other materials |

*From:*  Page MJ, McKenzie JE, Bossuyt PM, Boutron I, Hoffmann TC, Mulrow CD, et al. The PRISMA 2020 statement: an updated guideline for reporting systematic reviews. BMJ 2021;372:n71. doi: 10.1136/bmj.n71. This work is licensed under CC BY 4.0. To view a copy of this license, visit <https://creativecommons.org/licenses/by/4.0/>
